# Simultaneous identification of viruses and viral variants with programmable DNA nanobait

**DOI:** 10.1101/2021.11.05.21265890

**Authors:** Filip Bošković, Jinbo Zhu, Ran Tivony, Alexander Ohmann, Kaikai Chen, Mohammed F. Alawami, Milan Đorđević, Niklas Ermann, Joana Pereira Dias, Michael Fairhead, Mark Howarth, Stephen Baker, Ulrich F. Keyser

## Abstract

Respiratory infections are the major cause of death from infectious disease worldwide. Multiplexed diagnostic approaches are essential as many respiratory viruses have indistinguishable symptoms. We created self-assembled DNA nanobait that can simultaneously identify multiple short RNA targets. The nanobait approach relies on specific target selection via toehold-mediated strand displacement and rapid read-out via nanopore sensing. Here, we show this platform can concurrently identify several common respiratory viruses, detecting a panel of short targets of viral nucleic acids from multiple viruses. Our nanobait can be easily reprogrammed to discriminate viral variants, as we demonstrated for several key SARS-CoV-2 variants with single-nucleotide resolution. Lastly, we show that nanobait discriminates between samples extracted from oropharyngeal swabs from negative and positive SARS-CoV-2 patients without pre-amplification. Our system allows for multiplexed identification of native RNA molecules, providing a new scalable approach for diagnostics of multiple respiratory viruses in a single assay.

## Introduction

The diagnosis of infectious diseases plays a vital role in determining appropriate patient treatment^1^. Respiratory tract infections are the major cause of death from infectious diseases globally^2,3^. Many respiratory viruses induce comparable symptoms and cannot be differentiated clinically, making the identification of appropriate treatment challenging. It is estimated that 65 % of infection-associated cases of pneumonia are potentially misdiagnosed, with 95 % of these cases erroneously receiving antimicrobials^4^. The ongoing COVID-19 pandemic further highlights another unmet diagnostic need: the routine identification and screening of viral variants as they arise^5^.

Currently, viral diagnostics rely on quantitative reverse transcription-polymerase chain reaction (qRT-PCR), followed by genome sequencing to detect viral variants^5,6^. PCR-based diagnostic methods provide a sensitive approach for detecting viral nucleic acids in complex biological samples but suffer from limited multiplexing capabilities^7^. There is a need for robust diagnostic methods that can simultaneously detect multiple respiratory viruses and variants in limited sample volume, which can be quickly reconfigured to detect additional variants as they arise. Newer nucleic acid detection methods, such as nanopore sensing, which can distinguish multiple nucleic acid species^8–11^ with a unique signature for each designed DNA nanostructure may be an alternative approach for multiplexed biosensing^19,29,30^. Various groups have shown that nanopore sensing after viral nucleic acid enrichment or amplification may be a suitable platform to challenge these diagnostics^10,12,13^.

Here, aiming to solve many of the limitations for diagnostic multiplexing, we developed an innovative method that employs bespoke nanobait for the simultaneous identification of multiple respiratory viruses and variants^14^. We employed programmable viral RNA cutting with RNase H to remove short RNA targets that uniquely identifies the virus. The resultant RNA target is captured by nanobait, which is detected immediately by nanopore sensing, without reverse transcription, pre-amplification, or purification. By multiplexing several targets from the same virus in samples containing human RNA, we show that our method can increase specificity and throughput compared to existing platforms and paves the way for amplification-free RNA identification and diagnostics.

## Main text

### Single-molecule target RNA detection with nanobaits

We developed a workflow for nanobait detection of target RNA, ranging from patient swabbing, nucleic acid extraction, and programmable RNase H cutting of viral RNA (Figure 1). The RNA targets are selected by guide oligonucleotides (single-stranded DNA, 20 nt) that were designed to bind upstream and downstream on the specific regions in a viral genome. Then, RNase H was used to digest the RNA sequence in RNA: DNA hybrids (DNA guide oligo hybridized to viral RNA segment) and release the middle target RNA (Figure 1a, right).

**Figure 1.**
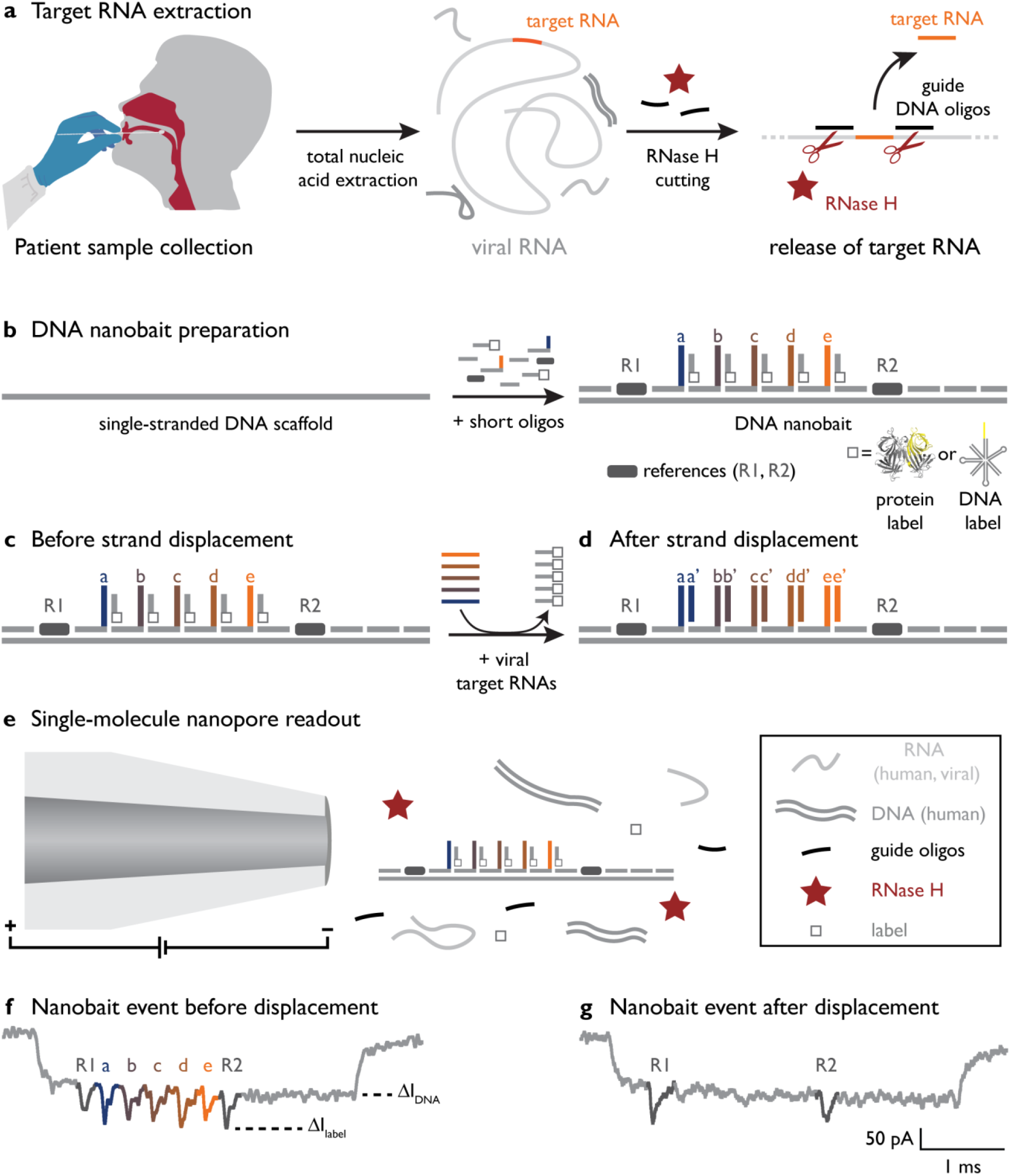
Self-assembled DNA nanobait strategy for multiplexed viral diagnostics. a) Oropharyngeal swab sample is collected from patients suspected to have COVID-19. Total nucleic acids were extracted (human and viral RNAs are shown in light gray; DNA is shown in dark gray), and target RNA was cleaved out using programmable RNase H cutting. Such treated sample was further tested for viral presence. b) Nanobait is made by mixing a single-stranded DNA scaffold (M13mp18 DNA) with short oligonucleotides where some of them carry complementary capture strands (a-e) for specific targets (a’-e’) in a target viral RNA. In addition, a partially complementary oligo with a structural label (protein or DNA-based, white square) is added for each site to amplify the signal in nanopore recordings. We marked a sensing region with two references (dark gray). Nanobait before c) and after d) strand displacement reactions of five targets (colored strands). If targets are present the five gray strands with labels are displaced. Two outer signals originate from reference structures that indicate the sensing region, and the five binding sites between the references are specific for the five different targets. e) Each nanobait is voltage-driven through the nanopore and detected in a mixture of molecules. f) Typical nanopore current signature as a function of time for a nanobait as designed with five labels present. The first current drop corresponds to DNA (ΔI_DNA_) and the second to labels (ΔI_label_). g) Typical nanobait event after strand displacement of all five targets. The presence of the targets is detected by the missing downward signals specific to each target.

Released RNA targets were identified using sequence specific binding to the nanobait (Figure 1b-c). The nanobait was designed (more details in Supplementary Figure S1) with five binding sites that could incorporate up to five targets. Nanobait was assembled by mixing a single-stranded DNA scaffold (linearized M13mp18, 7228 nt long)^19^ with a collection of short complementary oligonucleotides (Figure 1b; Figure S1; Supplementary Table S2). Towards one end of the nanobait, the sensing region was designed to contain equally spaced sites a-e flanked by two reference structures R1 and R2, which consisted of six DNA dumbbells each (oligonucleotides are listed in Supplementary Table S3). The sensing site contained a DNA overhang, which was fully complementary to the respective target sequence. We additionally exploited a blocking oligo with a label (monovalent streptavidin^20^ or DNA flower; Supplementary Figure S2 and Table S1) that was only partially hybridized and left six bases unpaired. Assembly of the nanobait was confirmed by AFM imaging and an electrophoretic mobility shift assay (EMSA) (Supplementary Figures S3-S4 and Figure S5, respectively). Ultimately, if the target was present, it would bind to the six unpaired bases and displace the blocking oligo with the label at its complementary overhang, which is known as toehold-mediated strand displacement^23^. Hence, the presence of the predefined targets was indicated by the absence of a label at the respective site (Figure 1c and Figure 1d).

We determined the structure of each nanobait and ability to detect the presence or absence of targets by a single-molecule readout technique exploiting nanopore resistive-pulse sensing (Figure 1e-g). Nanopore DNA sensing works via voltage-driven translocation of negatively charged nanobaits through a small orifice towards a positively charged electrode in an electrolyte solution (Figure 1e)^24^. Here, the nanobait translocation induces a unique current blockage signature (Figure 1f). The first current drop corresponded with double-stranded DNA nanobait (ΔI_DNA_). The second current drop (ΔI_label_) indicated the presence of the reference ‘R1, R2’ and labels ‘a-e’ (Figure 1c). Figure 1f depicts an exemplar nanobait nanopore event with seven downward spikes, where each spike corresponds with the matching color site on the schematic representation in Figure 1c. After strand displacement with all five targets present (a’-e’, Figure 1d), the five labeled oligos were displaced and only the reference spikes remain (Figure 1g). The short duplexes were significantly smaller than the labels and not detected with these nanopores^18^. Each ionic current event on a single nanobait revealed the presence of multiple short RNA targets. The flexibility of the nanobait design permitted us to identify targets originating from multiple parts of the same virus or from multiple viral genomes.

### Simultaneous detection of multiple viral variants

We designed nanobait for the multiplexed target identification of SARS-CoV-2, RSV (universal for group A), rhinovirus (universal), influenza (universal for group A), and parainfluenza 1 (Supporting information Table S4-S7). A schematical design of the nanobait for multiple respiratory viral nucleic acid targets is shown in Figure 2a. RSV is provided as an example of site-specific displacement (Figure 2a). The five targets, as well as the control (no target), were detected independently using the same nanobait. The first nanopore translocation events of the nanobait in each of the individual samples are depicted in Figure 2a and Supplementary Figure S6. Nanopore events with seven spikes indicated the absence of targets. If the respective target for SARS-CoV-2, RSV, rhinovirus, influenza, or parainfluenza were present, that spike was absent in the nanobait translocation event (presence of the targets are shown in Supplementary Table S4). The displacement efficiency was calculated as the difference between a no target control and the measurement for each site (fifty nanobait events for each of three nanopore recordings) (\*\*\**p*<0.001; two-sided Student’s T test). (Figure 2c). We tested two different scenarios, with and without targets for statistical significance.

**Figure 2.**
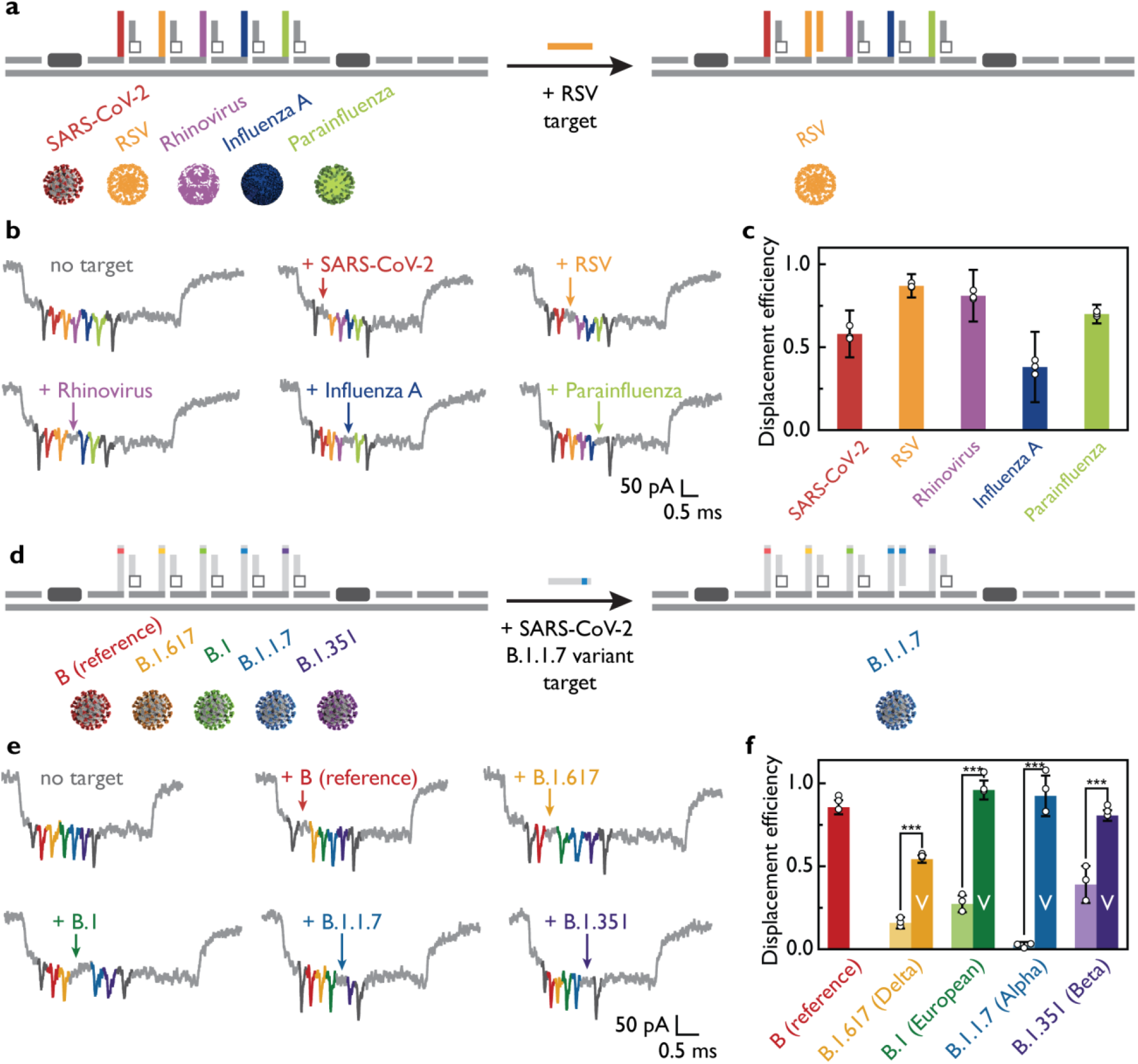
Multiplexed discrimination of viruses and SARS-CoV-2 variants with nanobait. a) Nanobait is designed to have five sites specific to SARS-CoV-2, RSV, rhinovirus, influenza A, and parainfluenza. Example events for the condition without any targets and for each virus-specific target are depicted in b). The absence of the colored spike indicates the presence of each respective target. c) Displacement efficiency indicates presence of corresponding viral target. The displacement efficiency represents a measurement with target subtracted from the control (no targets). Error bars represent standard error and the center as the mean for three nanopore measurements and fifty nanopore events per measurement. d) Nanobait designed to detect four single-nucleotide SARS-CoV-2 variants by adaptation of the target sequences. e) Example events for the condition without any targets and for each variant-specific target are depicted. The absence of the colored spike indicates the presence of each respective variant. f) Displacement efficiencies for single-nucleotide variants (labelled as ‘V’) are compared with the displacement efficiency for the wild-type Wuhan strain of the SARS-CoV-2 virus. Error bars represent standard error and the center as the mean for three nanopore measurements and fifty nanopore events per measurement. The difference between conditions without and with variant targets is statistically significant (***p < 0.001; two-sided Student’s T test; N=150).

Variant discrimination with single-nucleotide resolution is an essential feature for variant diagnostics. We tested the potential of nanobait for single nucleotide variant (SNV) discrimination, by distinguishing nucleic acids from several SARS-CoV-2 variants. The five sites of nanobait allowed for the simultaneous detection of wild-type virus and four variants (sequences are listed in Table 8-12, and design principles are elaborated in the Supporting information section S7)^25^. The first site was wild-type SARS-CoV-2 isolated in Wuhan (B by PANGOLIN nomenclature)^25^. The alternative four targets were European strain B.1 and three variants of concern:^14^ B.1.1.7 (alpha), B.1.351 (beta), and B.1.617 (delta) that were first detected in the United Kingdom, South Africa, and India, respectively. As an example, we highlight the identification of the B.1.1.7 variant (Figure 2d). We selected a variant-specific target that was fully complementary to the capture strand on the nanobait, while the wild type (WT) target contained a mismatch in the toehold end (Supplementary Table S11). The displacement efficiency is dependent on the number and position of mismatches in the toehold domain^26^. Programming nanobait with a single-nucleotide mismatch allowed us to discriminate the SARS-CoV-2 variant from the WT sequence. We depict example events for each sample where all spikes are present (no targets) or the respective spike is absent depending on which variant is present (Figure 2e; more events are shown in Supplementary Figure S7; presence of the targets are shown in Supplementary Table S8). Figure 2f shows the displacement efficiency for WT targets and their corresponding variant targets for the first fifty nanobait events (colored bars). We observed a significant difference for all four variants when compared to the respective WT samples (light and dark-colored bars). In addition, we demonstrated the principle by using two single-nucleotide SARS-CoV-2 RNA viral variants as shown in Figure S8 (more details in Section S8 of the Supplementary Information).

### Identification of multiple SARS-CoV-2 targets

Diagnostic tests for viral RNA rely on multistep reactions and subsequent purification steps. We aimed to use nanobait for direct target identification without preamplification and purification. Here, we used nanobait for specific single-molecule detection in a complex human transcriptome mixture i.e. human total universal RNA (htRNA; Invitrogen). These nanobait could identify multiple samples from pooled samples in complex backgrounds by nanopore sensing (Figure 3a).

**Figure 3.**
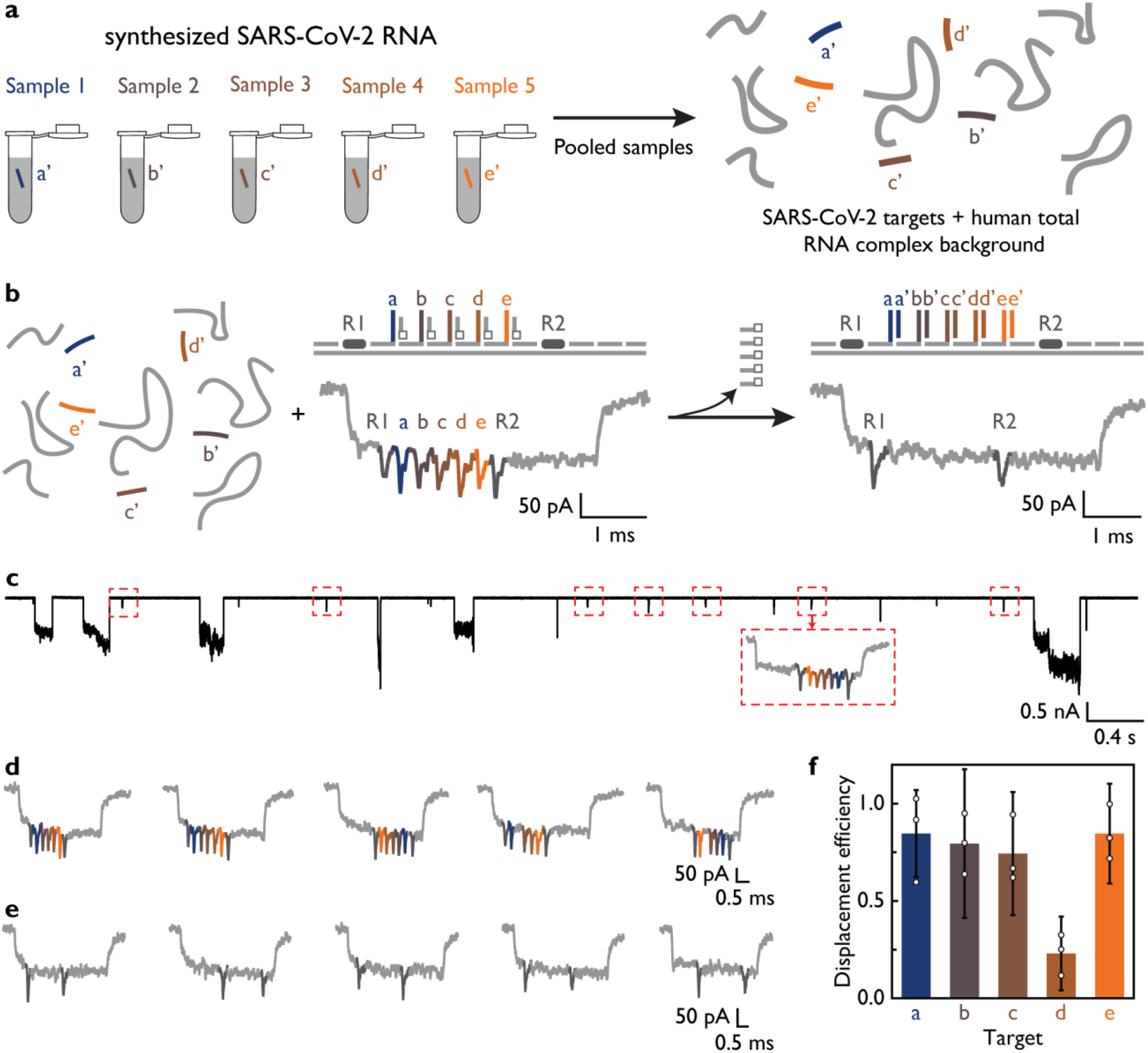
Nanobait detects multiple synthesized SARS-CoV-2 RNA targets in the background of the human transcriptional RNA. a) Five targets from different regions in a viral genome can be separately targeted and pooled for nanopore analysis. Five targets are mixed with intact human total RNA in the background, to verify that viral RNA purification is not a required step. b) After the addition of nanobait to a mix, all five targets can be identified in parallel as shown in the example events. c) Ionic current traces indicate the specificity of the method for the identification of nanobait-specific events even in a complex background where large downward signals originate from background including long RNAs. All nanobait events have been highlighted in red dashed boxes. d) First five single-file nanobait events for sample mixed with only human total RNA indicate correct current signature. e) The first five single-file nanobait events that have been previously mixed with targets and human total RNA. All targets are present since corresponding spikes are absent in nanopore events. f) Displacement efficiency calculated for the sample with targets added (nanobait with targets and human total RNA) for all five sites. Error bars represent standard error and the center as the mean for three nanopore measurements and fifty nanopore events per measurement.

We pooled five synthesized SARS-CoV-2 RNA targets together to investigate the specificity and potential crosstalk between nanobait and non-specific htRNA background. After the targets were added, all sites were displaced and correctly identified using the nanopore measurements (Figure 3b). A typical current trace indicates that nanobait spikes can be easily distinguished (red boxes, Figure 3c) from non-specific current blockages originating from the htRNA. Figures 3d and 3e show the first five linear nanobait events for samples with and without targets and in the presence of htRNA; the displacement efficiency for all five targets is depicted in Figure 3f. Target 4 had the lowest displacement efficiency, which was in agreement with a low predicted GC content of 25 %^27^. Nanobait based strand displacement can operate effectively even in a complex background of htRNA, oligonucleotides, and proteins. We studied the kinetic details for both RNA and DNA targets and determined that 10 minutes was the optimal incubation time for the strand displacement reaction (corresponding plots and events are presented in Supplementary section S9; presence of the targets are shown in Supplementary Table S14; target sequences and oligonucleotides are listed in Tables S15-S17).

### Design of target sites depends on viral RNA secondary structure

We next aimed to optimize multiple parameters in designing an efficient target RNA identification system. One key parameter was the successful excision of the short RNA targets from viral RNA. We found that the location of the target RNA in the viral RNA secondary structure affected the concentration of free target RNA and consequently affected displacement efficiency. A target in a highly complementary region would remain bound to the viral RNA after cutting and prevent detection. In contrast, the release of the target after RNase H excision increases when more unpaired bases were in the target region than within the secondary structure of the viral RNA. For future experiments, we maximized the number of unpaired bases to increase the effective concentration of the target in solution and consequently aid detection.

The role of unpaired bases was demonstrated by the detection of three targets in ∼3.6 kb RNA genome of MS2 virus (Figure 4a, the minimal free energy secondary structure^28^). The three targets (T1, T2, and T3) had a decreasing percentage of unpaired bases (T1 - 55%, T2 - 30%, and T3 - 25%). Subsequently, we designed oligos and employed RNase H cutting of all three targets and quantified the displacement efficiency using nanobait with the three sites (Figure 4b, more events are shown in Supplementary Figure S15; presence of the targets are shown in Supplementary Table S18; oligonucleotides are listed in Supplementary Tables S19-S23). Efficient cutting of viral RNA was confirmed by agarose gel electrophoresis (Supplementary Figure S11). For each target, the original 3.6 kb RNA was cut in fragments of the predicted length; additionally, the predicted fragment lengths were comparable when all three targets were cut simultaneously. We confirmed that target T1 was free in solution by hybridizing it to the complementary capture strand C and detected it using polyacrylamide gel electrophoresis (PAGE) (Figure 4c). After cutting, target T2 was not visible by PAGE (Supplementary Figure S14), and the oligonucleotides’ running speed and non-specific interactions were validated on a control PAGE (Supplementary Figure S12 and S13).

**Figure 4.**
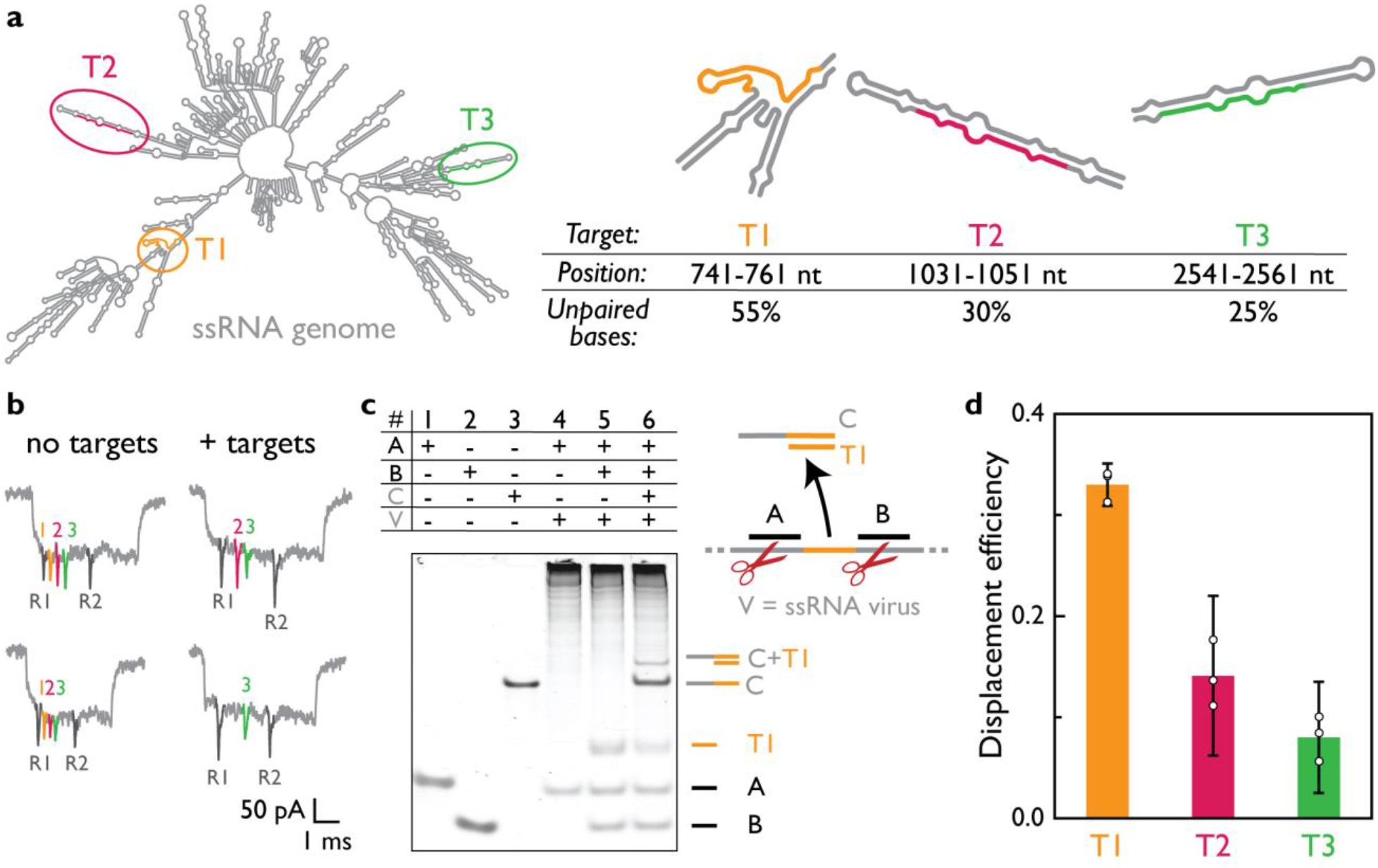
Position of a target in a viral RNA secondary structure influences the efficiency of target identification. a) We demonstrated that the position of a target in a viral RNA correlates with the efficiency of release of the target after RNase H cutting. MS2 viral RNA is presented as the minimal free energy structure. Three targets (T1, T2, T3) are selected to have a different level of paired bases for a constant 20 nt long target. For each target, a region in MS2 viral RNA is indicated with the percentage of unpaired bases in each target. b) Nanobait events with three targets present are shown, indicating the correct design (no targets). Events for the sample where cut long RNA is mixed with nanobait show displacement (+ targets). Each spike color corresponds to a site on nanobait. c) EMSA shows guide oligos A and B (lane 1 and 2, respectively), and complementary oligo C to target T1 (lane 3). If only guide oligo A is used for viral RNA cutting (lane 4), there is only the oligo A band and the high molecular weight cut viral RNA. Once both guide oligos are added, an additional band originating from released target T1 RNA emerges (lane 5). Once strand C is added to the same sample, we can see its shift after T1 binding (lane 6). d) Displacement efficiency of target RNAs correlates with the percentage of unpaired bases in a target. T1 has shown the highest displacement while T2 and T3 have lower displacement efficiencies. Error bars represent standard error and the center as the mean for three nanopore measurements and fifty nanopore events per measurement.

Example nanopore events and displacement efficiency with and without targets released from the MS2 RNA genome after RNase H cutting are shown in Figures 4b and 4d. The plot indicates that displacement was detected for all three targets. Target T1 had the highest displacement efficiency, while target T3 had the lowest displacement efficiency. As predicted, displacement efficiency (Figure 4d) correlated with unpaired base percentage in the RNA structure for each target, signifying an important design principle in selecting viral target regions for detection.

### Amplification-free SARS-CoV-2 identification in clinical samples

After establishing that RNase H had cut the MS2 RNA, we considered that nanobait could detect SARS-CoV-2 RNA in clinical samples. We accessed oropharyngeal swabs from patients suspected to have COVID-19; the viral load SARS-CoV-2 in oropharyngeal swabs in the clinical phase can be up to 10^8^ - 10^11^ copies^29,30^. The sensitivity curve for nanopore readout was plotted in Section S16, Figure S19. We used nanobait in nucleic acid extractions from clinical samples that had been prepared for qRT-PCR (more details in Supplementary section S12; oligos are listed in Supplementary Tables S24-S27)^15^. SARS-CoV-2 targets (S1, S2, and S3) were designed in conserved regions of the genome that contained the highest percentage of unpaired bases (Figure 5a). S1 was in the region encoding the spike (S) protein, S2 was in the region encoding the small envelope (E) glycoprotein, and S3 was in the nucleocapsid (N) protein-coding region. Total nucleic acids from clinical samples were subjected to our RNase H protocol (Figure 5b) and then mixed with a nanobait with sensing sites S1, S2, and S3. The reaction did not require further purification or preamplification before the nanopore readout. The nanobait mixture was then analyzed with nanopores containing the complex background of DNA (human and optionally DNA flower), long RNAs (human and potentially viral), short guide oligos, and proteins (RNase H and monovalent streptavidin).

**Figure 5.**
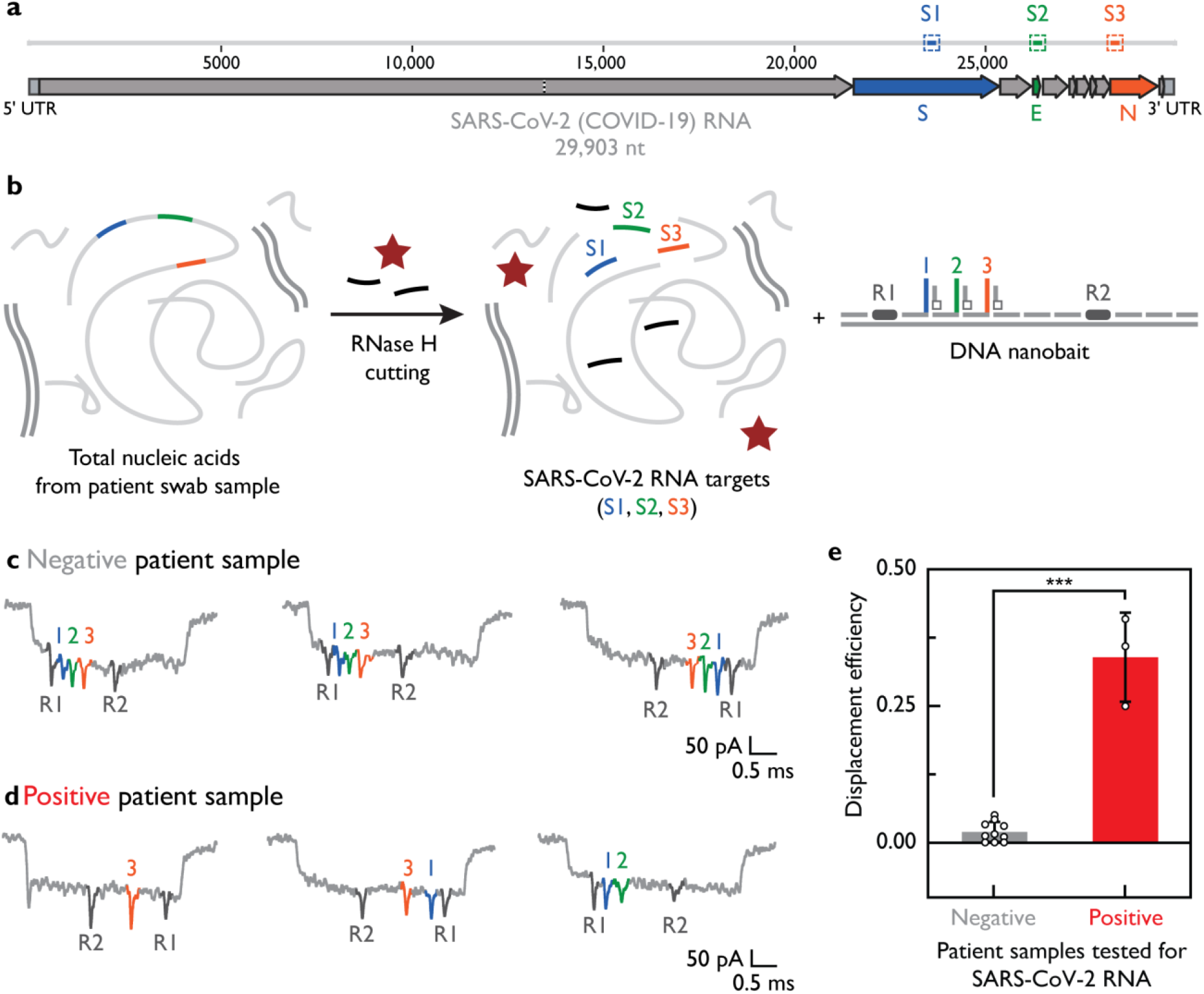
SARS-CoV-2 detection in patient oropharyngeal swab samples. a) We designed three targets (S1, S2, S3) in conserved regions that code for spike (S), envelope (E), and nucleocapsid (N) proteins as indicated in the schematic representation of the SARS-CoV-2 RNA genome (29,903 nt long). b) Total nucleic acids from oropharyngeal swabs contain a mix of human DNA and RNA that either have tested positive or negative for the COVID-19 with qRT-PCR. The next step included RNase H target release from long RNA and mixing with nanobait. If targets are present, they displace the oligo harboring a label. In this way, the displacement efficiency for each site is detectable with nanopores with 1 pM of nanobait. Real-time analysis in a mixture of various biomacromolecules (human DNA, human RNA, RNase H, streptavidin, guide oligos) is performed directly without prior purification enabling rapid nanopore readout (∼10 min). Example events for both negative and positive SARS-CoV-2 samples were presented in c) and d), respectively. e) The displacement efficiency in negative samples differs from positive samples. Error bars indicate standard error and the center as the mean for all events in the first ten minutes. Difference between negative and positive samples has statistical significance (***p < 0.001; two-sided Student’s T test). We used thirteen patient samples (N = 13).

Nanopore events from the nanobait mixed with RNase H treated negative patient samples (confirmed with qRT-PCR) are shown in Figure 5c. In addition to the two reference spikes (dark gray), three further spikes were visible and corresponded with sites for S1 (blue), S2 (green), and S3 (orange). As shown above, the nanobait current signature was not affected by the complex background or unspecific binding of DNA guide oligos. The missing spike associated with specific displacement was apparent when the nanobait was mixed with the SARS-CoV-2 positive swab samples as confirmed by qRT-PCR (Figure 5d). We repeated the procedure for in total thirteen SARS-CoV-2 clinical samples, which contained three positive and ten negative as shown by qRT-PCR. The nanobait displacement efficiencies for negative and positive samples were consistent with the qRT-PCR results (Figure 5e).

We additionally exploited a DNA flower as an alternative to a monovalent streptavidin using patient samples processed with RNase H cutting as well. We observed comparable results with this DNA-based system (more details in Supplementary section S13), indicating that the detection system can be based only on DNA. An all-DNA nanobait system may aid future upscaling.

Our nanobait approach bypasses pre-amplification and purification and hence avoids these potentially time-consuming and expensive steps. Furthermore, the nanopore readout time can be further reduced by performing real-time analysis on the QuipuNet convolutional neural network^21^. QuipuNet has high accuracy with an analysis speed of 1,600 events/s, which is more than sufficient for rapid viral detection. In this paper we employed standard RNA extraction procedures for qRT-PCR tests, the speed of test might further be improved by using simplified RNA extraction protocols or by combining it with RNase H cutting^31,32^.

## Conclusions

Here, we demonstrate the site-specific excision of a target from long viral RNA using RNase H cutting. In this way, we increase the displacement efficiency by ensuring the exact target sequence for displacement reaction in comparison to non-specific RNA fragmentation^33^. RNase H can be used to cut sequences next to a target sequence that yields new functionality besides its use in amplification-based viral detection protocols^34^. Additionally, site-specific RNA cutting can be achieved using DNAzymes or even the CRISPR/Cas system^35,36^.

Previous nanopore studies have demonstrated the ability to detect one or a limited number of short nucleic acid species in isolated form^10,12,33,37–39^. However, the biological complexity within a test sample possess a specific challenge when wanting to discriminate targets in this complex background^11,40^. Our work demonstrates that DNA nanotechnology can be used to detect specific targets in clinical samples with nanopores. As a proof of concept, we tested nanobait against five different respiratory viruses or SARS-CoV-2 variants in parallel. Previously, we showed that with DNA encoding a library of 2^112^ molecules that ensures the potential to test for 100s and 1000s of viral targets in parallel can be created^19,41^, especially when multiplexed nanopore systems become more advanced.

Recent studies have developed a viral nucleic acid detection system using nanopores which holds great promise for a rapid detection system^10,12,13^. However, preamplification and enzymatic steps in preparation for nanopore detection limit the utility of such methods, although some approaches showed potential to omit these steps^32,33,42^. Our nanobait system does not necessitate preamplification and can identify native RNA sequences without the need for sequencing. The design of this approach overcomes an issue of non-specific spikes in nanopore measurements by using the absence of a downward spike as a positive signal for identification of the presence of the target sequence. Nanobait demonstrated comparable features to the existing methods (Supplementary Data 1)^43–47^ and it can also identify multiple targets from the same viral RNA, hence offering enhanced specificity and accuracy for viral identification, as demonstrated for the detection of SARS-CoV-2 in clinical specimens. Currently, we show that 10 min nanobait readout with a nanopore would enable detection of viral RNA in infectious patients with Ct<20 (Figure 5). Nanopores have single-molecule sensitivity, however, the number of events depends on the target concentration^38^. Hence, lower concentrations (Ct>20) can be measured by either a single nanopore running for longer time or many nanopores in parallel. Here, the detection time scales with 1/N, where N is the number of nanopores.

Rapid programmability of diagnostic platforms is of paramount importance for detecting new viruses or their variants as they arise^14^. Nanobait is rapidly adaptable for new viral targets, as we demonstrated by discriminating the emerging SARS-CoV-2 variants. Our study has the potential to enable amplification-free native RNA identification. Nanobait bypasses amplification sequence biases by detecting innate RNA diversity. Our results show that nanobait identify short and long RNAs and may find wider applications in analysis of physiological and pathological conditions including cancer detection.

In conclusion, we have demonstrated the simultaneous identification of nucleic acids from multiple viruses and SARS-CoV-2 RNA variants in a specific and rapid manner by combining DNA nanotechnology and nanopore sensing. We employed the easily programmable nanobait with strand displacement for the discrimination between SARS-CoV-2 WT RNA from variant RNA, comprising three variants of epidemiological concern^14^. Finally, we successfully used the nanobait-based nanopore sensing method in clinical samples and could accurately determine the presence or absence of SARS-CoV-2 in patient swabs. Nanobait circumvents the need for reverse transcription, amplification, or reaction purification, therefore, bypassing enzymatic biases and some additional steps. As nanobait has proven to be specific and accurate for viral detection in patient samples, we think our platform can be employed for native RNA detection. Nanobait paves a way for a multiplexed amplification-free RNA detection method that is dependent only on the rapid single-molecule readout of the nanobait structure.

## Data Availability

All data produced in the present work are contained in the manuscript or supplementary information. Source data can be found in https://doi.org/10.17863/CAM.89753.

## Acknowledgments

We thank Prof Paul Lehner for his evaluation of the project and advice on unmet diagnostic needs and Dr Nicholas Bell for help with AFM imaging of DNA structures. We thank Tim Fitzmaurice for his operational help during the COVID-19 pandemic. We acknowledge the Statistics Clinic, Centre for Mathematical Sciences for their guidelines on a statistical analysis of data presented in this study.

## Author contributions

F.B. and U.F.K. conceived the idea. F.B., J.Z., K.C., and M.F.A. prepared nanobait designs. J.Z. and F.B. designed the sequences for respective targets. F.B. performed the experiments using monovalent streptavidin. J.Z. performed the experiments using DNA flower and assembled DNA flower. A.O. and F.B. performed gel electrophoresis and analyzed respective data. F.B. performed electrophoretic mobility shift assay. R.T. performed AFM imaging of nanobait. F.B., J.Z., and M.Đ. analyzed the nanopore data. J.P.D. and S.B. collected oropharyngeal patient samples, extracted nucleic acids, and performed qRT-PCR. M.F. and M.H. prepared monovalent streptavidin. F.B. and U.F.K. wrote the initial manuscript draft. F.B., J.Z., A.O. and K.C. prepared the draft manuscript. All authors contributed to the discussion and the final manuscript version.

## Methods

### Patient sample collection

Patient samples were collected by swabbing the back of the throat (oropharyngeal swab) of patients as previously described^15^. The samples were collected from patients with the COVID-19-like clinical picture and were tested with the qRT-PCR after nucleic acid extraction. Briefly, after collection, swabs were placed into a labeled sample tube containing lysis buffer (4 M guanidine thiocyanate, 25 mM Tris-HCl, 0.5% β-mercaptoethanol, and MS2 RNA (Roche; 200 ng/μl)). The tube was gently agitated to ensure the even distribution of lysis buffer. The safety steps have been previously described and performed in a certified CL2 laboratory^15^.

### Nucleic acid extraction

Total nucleic acid was extracted using spin column-based systems and as employed by standardized qRT-PCR testing^15^. The internal amplification control (MS2 (∼ 6 × 10^4^ PFU/ml) per 10 ml of lysis buffer) was added in the top-up lysis buffer (25 μl per 10 ml of lysis buffer). The sample was eluted in 100 μl of nuclease-free water (nfH_2_0, Invitrogen) and left to stand for 1 minute prior to centrifugation for 1 minute at 21,130 × g (15,000 rpm) in a benchtop microfuge. Eluted samples were directly subjected to qRT-PCR. The remaining nucleic acid extracts were stored at -80 °C and further used for nanobait-nanopore sensing.

### Quantitative reverse-transcription polymerase chain reaction (qRT-PCR) for SARS-CoV-2

SARS-CoV-2 detection was performed as previously described^15^. Per reaction, the master mix contained 12.5 μl of 2 × Luna Universal Probe One-Step reaction mix, 0.5 μl of 20 μM Wu forward primer (5’-ATGGGTTGGGATTATCCTAAATGTGA-3’), 0.5 μl of 20 μM Wu reverse primer (5’-GCAGTTGTGGCATCTCCTGATGAG-3’), 0.3 μl of 10 μM MGB Probe 3 fluorescein (5’-ATGCTTAGAATTATGGCCTCAC-3’), 0.5 μl of 10 μM of internal control forward primer for MS2 RNA, 0.5 μl of 10 μM internal control reverse primer for MS2 RNA, 0.3 μl of 10 μM internal probe (MS2 ROX), 1 μl of Luna WarmStart RT Enzyme Mix and 3.9 μl nfH_2_0. 20 μl of the master mix was aliquoted into each well of a 96-well plate and then combined with 5 μl of each extract. The MS2 internal extraction and amplification control that underwent the full extraction protocol was included as the negative extraction control in a minimum of two wells on the qRT-PCR plate. To determine potential contamination in the qRT-PCR process, 5 μl nfH_2_0 was included as the qRT-PCR negative control. 5 μl of spiked SARS-CoV-2 template plasmid was included in a single well as the qRT-PCR positive control. After adding 5 μl of each sample to its designated well, the plate was sealed with an optically clear plastic seal. The plate was centrifuged for 1 min at 2,000 × g (1,000 rpm) at 4 °C and then inserted in the qRT-PCR machine (QuantStudio, Thermo Fisher Scientific) and the run was parametrized. Signals for fluorescein (FAM) and carboxyrhodamine (ROX) were acquired. ROX was used to detect the internal MS2 control and FAM was used to detect SARS-CoV-2 RNA. The assay was performed for 2 minutes at 25 °C, 15 minutes at 50 °C (for the reverse-transcriptase), 2 minutes at 90 °C, before 45 cycles of 95 °C for 3 seconds followed by 60 °C for 30 seconds. Results were determined by confirmation of the correct positive controls (amplification of the plasmid), the extraction and amplification controls of all samples (ROX channel), no amplification in the negative controls, and consistent mean values of controls. SARS-CoV-2 positivity was confirmed by amplification in the FAM channel with an appropriate sigmoidal curve with a cycle threshold (CT) value of ≤36. The CT values of MS2 and MGB probe 3 were maintained to track the quality and reproducibility of the assay^16^.

### Programmable RNase H cutting for nanobait

For nanopore sensing, SARS-CoV-2 RNA controls, nucleic acid extracts (patient samples) or MS2 viral RNA were used further for the detection with nanobait. Firstly, we mixed guide oligos with the sample and heated it to 70 °C for 5 minutes. RNase H (5,000 units/ml, NEB) was added, mixed, and heated for 20 minutes at 37 °C to allow the enzyme to cut RNA in the DNA : RNA hybrid that effectively releases target RNA. RNase H was thermally inactivated by incubation at 65 °C for 10 min. Guide oligos were validated to not form intramolecular structures, homo-or heterodimers using NUPACK software^17^. For the measurement with the absent target, the same protocol including guide oligos was used. The control measurements show no displacement and hence we can exclude any substantial cross-binding from guide oligos.

### Viral target sequence properties for nanobait

The length of target, toehold length and GC content were selected to ensure optimal hybridization^18^. For the DNA nanobait designs, target sequences were selected to be in the conserved regions of a viral genome and had 40-60 % GC content to form a stable 20 bp duplex. The toehold length was selected to be 6 nt long and to have 40-60 % GC content. We tested all sequences for potential undesirable highly stable intramolecular interactions or homodimers using NUPACK software^17^. Then we performed a cross-reactivity check between multiple sites employed in each experiment^17^.

### Preparation of DNA flower for nanobait

We designed a DNA flower as another label for SARS-CoV-2 RNA detection from the patient samples. Three DNA flowers specific for each SARS-CoV-2 target (7-way junctions, 7WJa, 7WJb, and 7WJc) were prepared separately. Taking 7WJc as an example, 4 μM DNA strand J1, J2, J3, and J4c (Supplementary Table S1) were mixed together in TM buffer (10 mM Tris-HCl, 10 mM MgCl_2_, pH 8.0) and heated to 90 °C for 5 min, then cooled down to 65 °C for 15 min, 45 °C for 15 min, 37 °C for 20 min, 25 °C for 20 min and finally 4 °C for 20 min. Strand J4c was substituted by J4b to prepare 7WJb. For 7WJa, to avoid self-folding at site 43 on the nanobait, J1, J2, J3 J4a, and C43 were mixed together before annealing.

### Self-assembly of DNA nanobait

DNA nanobait was assembled by mixing linearized single-stranded M13 DNA (m13mp18, 7249 nt, Guild Biosciences, 100 nM) with short complementary oligonucleotides^19^ some of those harboring reference structures, capture strands, and by adding partially complementary strands that were 3’-biotinylated for toehold-mediated strand displacement reaction. The linearized M13 DNA (7228 nt in length) was complemented by oligonucleotides hence creating a nicked double-stranded nanobait with two terminal 4 dT overhangs that prevent multimerization^19^. The mix contained 20 nM of linearized M13 DNA, 60 nM of oligonucleotides (three times excess to M13 DNA), 3’-biotinylated strands in the concentration of 180 nM, 10 mM MgCl_2_, and 1 × TE (10 mM Tris-HCl, 1 mM EDTA, pH 8.0). It was mixed by pipetting and spun down before heating to 70 °C for 30 s and cooled down over 45 min to ambient temperature. Excess oligonucleotides were removed using Amicon® ultra 0.5 mL centrifugal filters with 100 kilodalton (kDa) cutoff with the washing buffer (10 mM Tris-HCl pH 8.0, 0.5 mM MgCl_2_). If DNA flowers were employed as a label, the partially complementary strands that carry it were incubated in 10 mM MgCl_2_ for 2 h at ambient temperature, and subsequently, Amicon filtration was performed as described above. The asymmetry of the nanobait design allows for unambiguous identification of the binding sites. Nanobait was stored until used for further experiments under 4-10 °C in 0.5 mM MgCl_2_, 10 mM Tris-HCl, pH 8.0. Nanobait design was checked by nanopore readout before each measurement.

### Nanopore readout of DNA nanobait

Nanobait was mixed with a sample (nucleic acid extract or purified viral targets at ten times excess) in 10 mM MgCl_2_ and 100 mM NaCl. The mixture (5 μL) was incubated at room temperature (∼10 min) until prepared for nanopore measurement. The difference in a target sequence composition and its physical characteristics might lead to the variability in hybridization and hence the displacement efficiency of sensing sites^18^. We have used human total universal RNA (htRNA, 100 ng/μL, Invitrogen) as a background where indicated, to show that there are no nonspecific signals induced by human native RNAs. For nanopore measurement, the sample was diluted to <0.5 nM nanobait (for purified viral targets) or 4.7 μL of RNase H cut patient sample was mixed with 0.3 μL of monovalent streptavidin (SAe1D3)^20^ (1 μM), 5 μL of LiCl (4 M), and 5 μL of LiCl (8 M). We have fabricated 14 ± 3 nm (mean ± s.d.) nanopores^19^ using quartz glass capillaries with 0.5 mm outer diameter and 0.2 inner diameter (Sutter Instrument) by laser-assisted puller P-2000 (Sutter Instrument). The mix was pipetted in a nanopore PDMS (polydimethylsiloxane) chip, and all measurements were performed at a constant voltage of 600 mV. Nanopore measurement details are shown in Supplementary Table S30.

### Real-time nanopore data analysis

Nanopore data analysis is explained in detail in Supplementary section S14. Briefly, nanobait events were filtered out of raw ionic current traces, then the detection region was determined, and information of the spike’s presence at each specific site was extracted. The plotted displacement efficiency was calculated as a displacement efficiency for a measurement subtracted to a no target control for each site (fifty nanobait events for each of three nanopore recordings) unless stated otherwise:

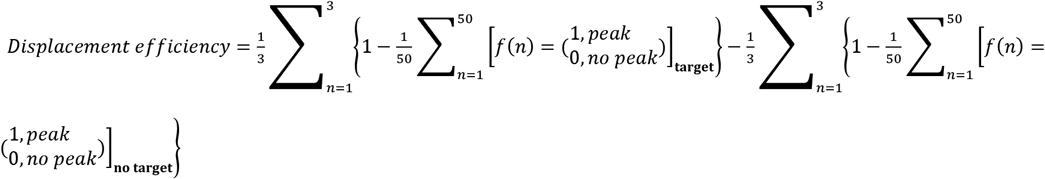

We verified that the convolutional neural network QuipuNet^21^ was capable of real-time analysis of nanopore data following the described procedure. Previously, we demonstrated that with around 10 events we reach 99 % confidence in a positive detection of our designed DNA structures^22^.

### AFM imaging

Atomic Force Microscopy (Nanosurf Mobile S) imaging of nanobaits was performed in air in non-contact mode. Nanobait structures were diluted to 1 ng/μL in 1 mM MgCl_2_ and 10 μL was added to freshly cleaved mica, incubated for 1 min, rinsed with filtered Milli-Q H_2_O, and then blow-dried with nitrogen. Prior to scanning, the mica plate was affixed to the AFM sample stage using double-sided adhesive tape. Image visualization and analysis were performed using Gwyddion.

### Statistical analysis

99.9% confidence intervals for the displacement efficiencies were calculated for all measurements. The statistical significance between two sites without and with the target was tested using a two-sided Student’s T-test.

## Data Availability

Data supporting the findings of this study are available in the main text and the Supplementary Information. Additional raw data are available at https://doi.org/10.17863/CAM.89753. Source data are provided with this paper.

