## Supplementary Information - materials and methods, figures and tables for "Simultaneous identification of viruses and viral variants with programmable DNA nanobait"

[*] *corresponding author*

This file includes:

Supplementary Text
Figs. S1 to S19
Tables S1 to S30
References

### Section S1. DNA nanobait

#### *Section S1.1 Nanobait design*

Nanobait is assembled by mixing the linearized DNA scaffold (M13mp18, 7228 nt, Guild Biosciences, 100 nM) with short complementary oligos[1]. The basic set of oligos has 188 oligos which are 38 nt long and two terminal oligos that are 46 nt long (with four terminal T to avoid aggregation by blunt-end stacking[1–3]) that are complementary to the 7228 nt long scaffold. These oligos are provided in Table S2. Certain oligos from Table S2 were replaced with oligos that code references (listed in Table S3) and capture oligos that are specific for five targets. Nanobait has a detection region that is marked by two references and contains five capture strands specific to five targets (Figure S1a).

Each reference has six interspaced DNA dumbbells (schematic design is shown in Figure S1b) i.e. DNA double hairpins (5'-TCCTCTTTTGAGGAACAAGTTTTCTTGT-3') that are interpreted as one downward signal in nanopore events due to their close proximity[4,5]. In addition to the capturing oligo (which contains two regions, one comprising 38 nt complementary to the DNA scaffold and one comprising 20 nt complementary to the target), each capture site contains a partially complementary oligo (14 nt) that carries a label (Figure S1c). This single-stranded region of the capture strand contains a toehold (6 nt) needed for strand displacement reaction (SDR). Once a target is added to the solution, it first binds to the toehold and then it's able to displace the oligo with a label (Figure S1d). Hence, target presence is detected as the absence of oligo with a label (which is read as a missing peak in nanopore event).

#### *Section S1.2 Linearization of circular single-stranded M13*

Linearization of circular M13 scaffold (M13mp18, 7249 nt) is performed using double restriction digestion. Firstly, we bound an oligonucleotide (39 nt long, 5'-TCTAGAGGATCCCCGGGTACCGAGCTCGAATTCGTAATC-3') to the scaffold to create a double-stranded region for efficient enzyme digestion. Such a double-stranded region contains closely spaced restriction sites for EcoRI and BamHI enzymes. The double enzyme digestion is used to ensure complete linearization of the scaffold. We mixed 52 µL of scaffold (M13mp18, 100 nM) with 10 µL of 10 × CutSmart buffer (1 × buffer components are 50 mM potassium acetate, 20 mM Tris-acetate, 10 mM magnesium acetate, 100 µg/ml BSA, pH 7.9 at 25 °C; New England Biolabs), 2.5 µL of 39 nt long oligo (100 µM in $H_20$, IDT), and 33.5 µL of filtered Milli-Q water. The mix was heated to 65 °C for 30 s and slowly cooled down to 25 °C over ~40 min. After oligo binding, 1 µL of EcoRI-HF enzyme (100,000 units/mL) and1 µL of BamHI-HF enzyme (100,000 units/mL) was added to the reaction and mixed by pipetting the full volume multiple times and incubated at 37 °C for 1 h. The cut scaffold was purified as previously described using NucleoSpin Gel and PCR Clean-up kit (Macherey-Nagel™)[1].

*Section S1.3 Nanobait assembly*

Nanobait is assembled by mixing the scaffold with a respective set of oligos. Briefly, we mixed 800 fmoles of the linearized scaffold with three times excess of oligos (2400 fmoles each) in 10 mM $MgCl_2$, 1 × Tris-HCl, pH 8.0. The reaction was heated to 70 °C for 30 s followed by linear cooling to 25 °C over 45 min. The excess oligos were removed with Amicon 0.5 mL filters with 100 kDa cut-off using a washing buffer (10 mM Tris-HCl, pH 8.0, 0.5 mM $MgCl_2$). The samples were centrifuged at 9,200 × g for 10 min twice, and after the filter was inverted, placed in a new tube, and spun down at 1,000 × g for 2 min. Nanobait concentration was estimated from a NanoDrop spectrophotometer or a Qubit fluorometer using Qubit™ dsDNA BR Assay Kit.

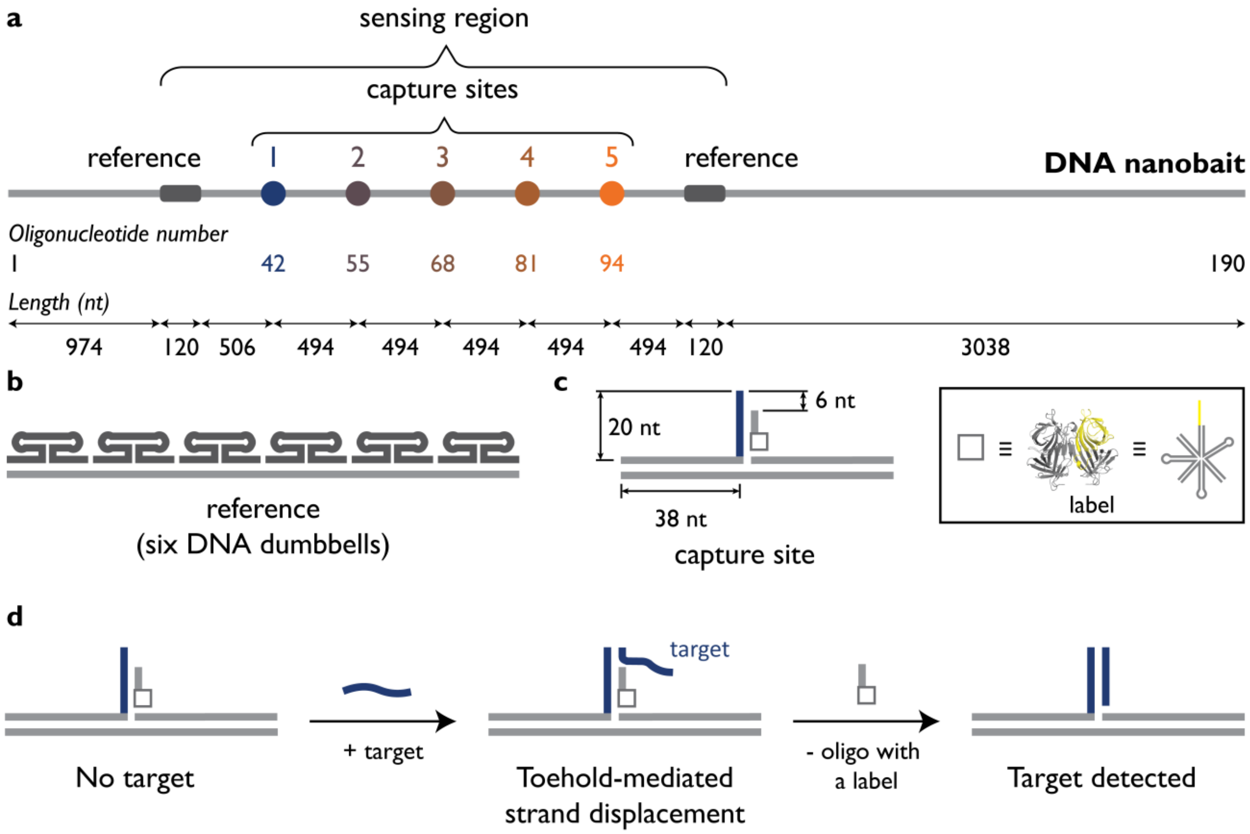

**Figure S1.** DNA nanobait schematic design. a) Nanobait contains a detection region that has two references (dark gray) and five capture sites for each target strand. Nanobait represents M13 DNA that is fully covered by oligonucleotides forming a double-stranded DNA with nicks. A standard set of 190 oligonucleotides was used and some of the oligonucleotides were replaced with a reference or capture strands. Oligo numbers replaced for each capture site is depicted below the nanobait. Lengths and distances between features on nanobait are represented as a number of nucleotides i.e. basepairs once a double-stranded molecule is formed. b) Reference contains six interspaced DNA dumbbells which are read as one downward signal in nanopore measurements. c) Capture site structure is represented by capture strand that has a 38 nt part complementary to the DNA scaffold and a 20 nt part that is fully complementary to a target strand. The oligo carrying the label is partially complementary to the capture strand (14 nt). The label can be either monovalent streptavidin or a DNA flower. d) The uncomplemented part of the capture strand (6 nt) is called the toehold and it serves as a seed for displacement (removal) of the oligo with a label by the target.

### Section S2. Design of structural labels used for nanopore signal amplification

Specific identification of the presence or absence of the target sequence in solution is done by using nanopore sensing. In nanopore recordings, the “control” or absence of a target is detected as a downward signal or peak on a DNA event. This peak corresponds to a label that is attached to an oligonucleotide (oligo or strand) (further explained in Section S3). In this study, we demonstrated that both protein-based and DNA-based structures can be employed as labels.

As a protein-based label, we employed monovalent streptavidin (structure shown in Figure S2a) that has a single femtomolar affinity biotin binding site. Monovalent streptavidin expression and purification have been described previously[6]. Oligos have a biotin label on the 3’ end via a C6 spacer arm (Integrated DNA Technologies) to which monovalent streptavidin binds.

As a DNA-based label, we assembled a DNA flower[7] that represents a 7-way junction as is illustrated in Figure S2b using four DNA strands as described in the next section. One of the strands (J4, Table S1) has 14 nt single-stranded part (in yellow) that is used as an anchor that binds to nanobait (further explained in Section S3).

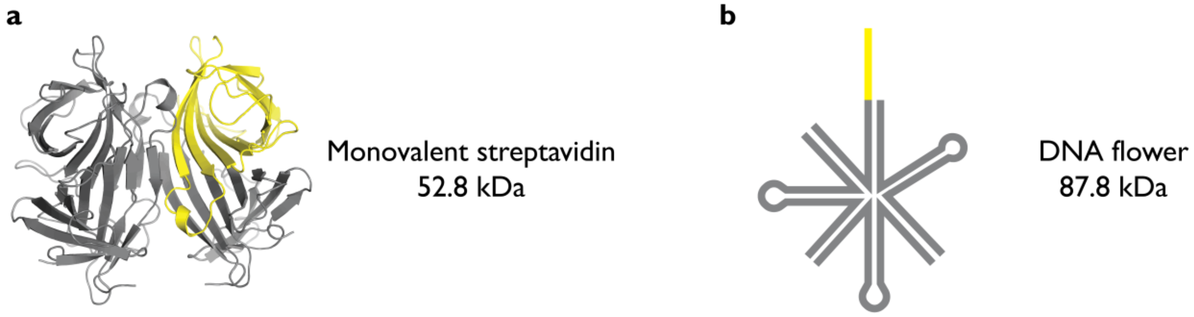

**Figure S2.** The designs of structures used in this study as labels. a) The tetramer is formed from one biotin-binding subunit in yellow and 3 non-binding subunits in gray (PDB ID: 5TO2). b) DNA flower represents a 7-way junction, designed with an arm length of 16 bp and loops with 4 T (gray). The region of oligo that binds to a respective site on the nanobait is in yellow.

### Section S3. DNA flower synthesis

Three DNA flowers specific for each SARS-CoV-2 target (7-way junctions, 7WJa, 7WJb, and 7WJc) were prepared separately. Taking 7WJc as an example, 4 µM DNA strands J1, J2, J3, and J4c (Table S1) were mixed in TM buffer (10 mM Tris-HCl, 10 mM $MgCl_2$, pH 8.0) and heated at 90 °C for 5 min, then cooled down to 65 °C for 15 min, 45 °C for 15 min, 37 °C for 20 min, 25 °C for 20 min and finally 4 °C for 20 min. Strand J4c was substituted by J4b to prepare 7WJb. For 7WJa, to avoid self-folding and improve the occupied fraction at site 43 on nanobait, J1, J2, J3 J4a, and C43 were mixed before annealing.

**Supplementary Table S1.** DNA flower oligonucleotides. The complementary parts are highlighted by using the same color.

| Strand name | Sequence (5'→'3') |
|---|---|
| J1 | GGATCAGAGCTGGACG ACAATGACGTAGGTCC TTTT GGACCTACGTCATTGT ACTATGGCACACATCC |
| J2 | GCAAGACTCGTGCTCA CCGAATGCCACCACGC TTTT GCGTGGTGGCATTCGG CGTCCAGCTCTGATCC |
| J3 | GGTTCAGCCGCAATCC TCGCCTGCACTCTACC TTTT GGTAGAGTGCAGGCGA TGAGCACGAGTCTTGC |
| J4a | TACTGCGCTTCGAT TT GGATGTGTGCCATAGT GGATTGCGGCTGAACC |
| J4b | AACTGAGGGAGCCT TT GGATGTGTGCCATAGT GGATTGCGGCTGAACC |
| J4c | AGACTAATTCTCCT TT GGATGTGTGCCATAGT GGATTGCGGCTGAACC |

**Supplementary Table S2.** DNA scaffold complementary oligonucleotides

| Oligo number | Sequence (5'→'3') |
|---|---|
| 1 | TTTTCGTAATCATGGTCATAGCTGTTTCCTGTGTGAAATTGTTATC |
| 2 | CGCTCACAATTCCACACAACATACGAGCCGGAAGCATA |
| 3 | AAGTGTAAAGCCTGGGGTGCCTAATGAGTGAGCTAACT |
| 4 | CACATTAATTGCGTTGCGCTCACTGCCCGCTTTCCAGT |
| 5 | CGGGAAACCTGTCGTGCCAGCTGCATTAATGAATCGGC |
| 6 | CAACGCGCGGGGAGAGGCGGTTTGCGTATTGGGCGCCA |
| 7 | GGGTGGTTTTTCTTTTCACCAGTGAGACGGGCAACAGC |
| 8 | TGATTGCCCTTCACCGCCTGGCCCTGAGAGAGTTGCAG |
| 9 | CAAGCGGTCCACGCTGGTTTGCCCCAGCAGGCGAAAAT |
| 10 | CCTGTTTGATGGTGGTTCCGAAATCGGCAAAATCCCTT |
| 11 | ATAAATCAAAAGAATAGCCCGAGATAGGGTTGAGTGTT |
| 12 | GTTCCAGTTTGGAACAAGAGTCCACTATTAAAGAACGT |
| 13 | GGACTCCAACGTCAAAGGGCGAAAAACCGTCTATCAGG |
| 14 | GCGATGGCCCACTACGTGAACCATCACCCAAATCAAGT |
| 15 | TTTTTGGGGTCGAGGTGCCGTAAAGCACTAAATCGGAA |
| 16 | CCCTAAAGGGAGCCCCCGATTTAGAGCTTGACGGGGAA |
| 17 | AGCCGGCGAACGTGGCGAGAAAGGAAGGGAAGAAAGCG |
| 18 | AAAGGAGCGGGCGCTAGGGCGCTGGCAAGTGTAGCGGT |
| 19 | CACGCTGCGCGTAACCACCACACCCGCCGCGCTTAATG |
| 20 | CGCCGCTACAGGGCGCGTACTATGGTTGCTTTGACGAG |
| 21 | CACGTATAACGTGCTTTCCTCGTTAGAATCAGAGCGGG |
| 22 | AGCTAAACAGGAGGCCGATTAAAGGGATTTTAGACAGG |
| 23 | AACGGTACGCCAGAATCCTGAGAAGTGTTTTTATAATC |
| 24 | AGTGAGGCCACCGAGTAAAAGAGTCTGTCCATCACGCA |
| 25 | AATTAACCGTTGTAGCAATACTTCTTTGATTAGTAATA |
| 26 | ACATCACTTGCCTGAGTAGAAGAACTCAAACTATCGGC |
| 27 | CTTGCTGGTAATATCCAGAACAATATTACCGCCAGCCA |
| 28 | TTGCAACAGGAAAAACGCTCATGGAAATACCTACATTT |
| 29 | TGACGCTCAATCGTCTGAAATGGATTATTTACATTGGC |
| 30 | AGATTCACCAGTCACACGACCAGTAATAAAAGGGACAT |
| 31 | TCTGGCCAACAGAGATAGAACCCTTCTGACCTGAAAGC |
| 32 | GTAAGAATACGTGGCACAGACAATATTTTTGAATGGCT |
| 33 | ATTAGTCTTTAATGCGCGAACTGATAGCCCTAAAACAT |
| 34 | CGCCATTAAAAATACCGAACGAACCACCAGCAGAAGAT |
| 35 | AAAACAGAGGTGAGGCGGTCAGTATTAACACCGCCTGC |
| 36 | AACAGTGCCACGCTGAGAGCCAGCAGCAAATGAAAAAT |
| 37 | CTAAAGCATCACCTTGCTGAACCTCAAATATCAAACCC |
| 38 | TCAATCAATATCTGGTCAGTTGGCAAATCAACAGTTGA |
| 39 | AAGGAATTGAGGAAGGTTATCTAAAATATCTTTAGGAG |
| 40 | CACTAACAACTAATAGATTAGAGCCGTCAATAGATAAT |
| 41 | ACATTTGAGGATTTAGAAGTATTAGACTTTACAAACAA |
| 42 | TTCGACAACTCGTATTAAATCCTTTGCCCGAACGTTAT |
| 43 | TAATTTTAAAAGTTTGAGTAACATTATCATTTTGCGGA |

| | |
|---|---|
| 44 | ACAAAGAAACCACCAGAAGGAGCGGAATTATCATCATA |
| 45 | TTCCTGATTATCAGATGATGGCAATTCATCAATATAAT |
| 46 | CCTGATTGTTTGGATTATACTTCTGAATAATGGAAGGG |
| 47 | TTAGAACCTACCATATCAAAATTATTTGCACGTAAAAC |
| 48 | AGAAATAAAGAAATTGCGTAGATTTTCAGGTTTAACGT |
| 49 | CAGATGAATATACAGTAACAGTACCTTTTACATCGGGA |
| 50 | GAAACAATAACGGATTCGCCTGATTGCTTTGAATACCA |
| 51 | AGTTACAAAATCGCGCAGAGGCGAATTATTCATTTCAA |
| 52 | TTACCTGAGCAAAAGAAGATGATGAAACAAACATCAAG |
| 53 | AAAACAAAATTAATTACATTTAACAATTTCATTTGAAT |
| 54 | TACCTTTTTTAATGGAAACAGTACATAAATCAATATAT |
| 55 | GTGAGTGAATAACCTTGCTTCTGTAAATCGTCGCTATT |
| 56 | AATTAATTTTCCCTTAGAATCCTTGAAAACATAGCGAT |
| 57 | AGCTTAGATTAAGACGCTGAGAAGAGTCAATAGTGAAT |
| 58 | TTATCAAAATCATAGGTCTGAGAGACTACCTTTTTAAC |
| 59 | CTCCGGCTTAGGTTGGGTTATATAACTATATGTAAATG |
| 60 | CTGATGCAAATCCAATCGCAAGACAAAGAACGCGAGAA |
| 61 | AACTTTTTCAAATATATTTTAGTTAATTTCATCTTCTG |
| 62 | ACCTAAATTTAATGGTTTGAAATACCGACCGTGTGATA |
| 63 | AATAAGGCGTTAAATAAGAATAAACACCGGAATCATAA |
| 64 | TTACTAGAAAAAGCCTGTTTAGTATCATATGCGTTATA |
| 65 | CAAATTCTTACCAGTATAAAGCCAACGCTCAACAGTAG |
| 66 | GGCTTAATTGAGAATCGCCATATTTAACAACGCCAACA |
| 67 | TGTAATTTAGGCAGAGGCATTTTCGAGCCAGTAATAAG |
| 68 | AGAATATAAAGTACCGACAAAAGGTAAAGTAATTCTGT |
| 69 | CCAGACGACGACAATAAACAACATGTTCAGCTAATGCA |
| 70 | GAACGCGCCTGTTTATCAACAATAGATAAGTCCTGAAC |
| 71 | AAGAAAAATAATATCCCATCCTAATTTACGAGCATGTA |
| 72 | GAAACCAATCAATAATCGGCTGTCTTTCCTTATCATTC |
| 73 | CAAGAACGGGTATTAAACCAAGTACCGCACTCATCGAG |
| 74 | AACAAGCAAGCCGTTTTTATTTTCATCGTAGGAATCAT |
| 75 | TACCGCGCCCAATAGCAAGCAAATCAGATATAGAAGGC |
| 76 | TTATCCGGTATTCTAAGAACGCGAGGCGTTTTAGCGAA |
| 77 | CCTCCCGACTTGCGGGAGGTTTTGAAGCCTTAAATCAA |
| 78 | GATTAGTTGCTATTTTGCACCCAGCTACAATTTTATCC |
| 79 | TGAATCTTACCAACGCTAACGAGCGTCTTTCCAGAGCC |
| 80 | TAATTTGCCAGTTACAAAATAAACAGCCATATTATTTA |
| 81 | TCCCAATCCAAATAAGAAACGATTTTTTGTTTAACGTC |
| 82 | AAAAATGAAAATAGCAGCCTTTACAGAGAGAATAACAT |
| 83 | AAAAACAGGGAAGCGCATTAGACGGGAGAATTAACTGA |
| 84 | ACACCCTGAACAAAGTCAGAGGGTAATTGAGCGCTAAT |
| 85 | ATCAGAGAGATAACCCACAAGAATTGAGTTAAGCCCAA |
| 86 | TAATAAGAGCAAGAAACAATGAAATAGCAATAGCTATC |
| 87 | TTACCGAAGCCCTTTTTAAGAAAAGTAAGCAGATAGCC |
| 88 | GAACAAAGTTACCAGAAGGAAACCGAGGAAACGCAATA |
| 89 | ATAACGGAATACCCAAAAGAACTGGCATGATTAAGACT |
| 90 | CCTTATTACGCAGTATGTTAGCAAACGTAGAAAATACA |

| | |
|---|---|
| 91 | TACATAAAGGTGGCAACATATAAAAGAAACGCAAAGAC |
| 92 | ACCACGGAATAAGTTTATTTTGTCACAATCAATAGAAA |
| 93 | ATTCATATGGTTTACCAGCGCCAAAGACAAAAGGGCGA |
| 94 | CATTCAACCGATTGAGGGAGGGAAGGTAAATATTGACG |
| 95 | GAAATTATTCATTAAAGGTGAATTATCACCGTCACCGA |
| 96 | CTTGAGCCATTTGGGAATTAGAGCCAGCAAAATCACCA |
| 97 | GTAGCACCATTACCATTAGCAAGGCCGGAAACGTCACC |
| 98 | AATGAAACCATCGATAGCAGCACCGTAATCAGTAGCGA |
| 99 | CAGAATCAAGTTTGCCTTTAGCGTCAGACTGTAGCGCG |
| 100 | TTTTCATCGGCATTTTCGGTCATAGCCCCCTTATTAGC |
| 101 | GTTTGCCATCTTTTCATAATCAAAATCACCGGAACCAG |
| 102 | AGCCACCACCGGAACCGCCTCCCTCAGAGCCGCCACCC |
| 103 | TCAGAACCGCCACCCTCAGAGCCACCACCCTCAGAGCC |
| 104 | GCCACCAGAACCACCACCAGAGCCGCCGCCAGCATTGA |
| 105 | CAGGAGGTTGAGGCAGGTCAGACGATTGGCCTTGATAT |
| 106 | TCACAAACAAATAAATCCTCATTAAAGCCAGAATGGAA |
| 107 | AGCGCAGTCTCTGAATTTACCGTTCCAGTAAGCGTCAT |
| 108 | ACATGGCTTTTGATGATACAGGAGTGTACTGGTAATAA |
| 109 | GTTTTAACGGGGTCAGTGCCTTGAGTAACAGTGCCCGT |
| 110 | ATAAACAGTTAATGCCCCCTGCCTATTTCGGAACCTAT |
| 111 | TATTCTGAAACATGAAAGTATTAAGAGGCTGAGACTCC |
| 112 | TCAAGAGAAGGATTAGGATTAGCGGGGTTTTGCTCAGT |
| 113 | ACCAGGCGGATAAGTGCCGTCGAGAGGGTTGATATAAG |
| 114 | TATAGCCCGGAATAGGTGTATCACCGTACTCAGGAGGT |
| 115 | TTAGTACCGCCACCCTCAGAACCGCCACCCTCAGAACC |
| 116 | GCCACCCTCAGAGCCACCACCCTCATTTTCAGGGATAG |
| 117 | CAAGCCCAATAGGAACCCATGTACCGTAACACTGAGTT |
| 118 | TCGTCACCAGTACAAACTACAACGCCTGTAGCATTCCA |
| 119 | CAGACAGCCCTCATAGTTAGCGTAACGATCTAAAGTTT |
| 120 | TGTCGTCTTTCCAGACGTTAGTAAATGAATTTTCTGTA |
| 121 | TGGGATTTTGCTAAACAACTTTCAACAGTTTCAGCGGA |
| 122 | GTGAGAATAGAAAGGAACAACTAAAGGAATTGCGAATA |
| 123 | ATAATTTTTTCACGTTGAAAATCTCCAAAAAAAAGGCT |
| 124 | CCAAAAGGAGCCTTTAATTGTATCGGTTTATCAGCTTG |
| 125 | CTTTCGAGGTGAATTTCTTAAACAGCTTGATACCGATA |
| 126 | GTTGCGCCGACAATGACAACAACCATCGCCCACGCATA |
| 127 | ACCGATATATTCGGTCGCTGAGGCTTGCAGGGAGTTAA |
| 128 | AGGCCGCTTTTGCGGGATCGTCACCCTCAGCAGCGAAA |
| 129 | GACAGCATCGGAACGAGGGTAGCAACGGCTACAGAGGC |
| 130 | TTTGAGGACTAAAGACTTTTTCATGAGGAAGTTTCCAT |
| 131 | TAAACGGGTAAAATACGTAATGCCACTACGAAGGCACC |
| 132 | AACCTAAAACGAAAGAGGCAAAAGAATACACTAAAACA |
| 133 | CTCATCTTTGACCCCCAGCGATTATACCAAGCGCGAAA |
| 134 | CAAAGTACAACGGAGATTTGTATCATCGCCTGATAAAT |
| 135 | TGTGTCGAAATCCGCGACCTGCTCCATGTTACTTAGCC |
| 136 | GGAACGAGGCGCAGACGGTCAATCATAAGGGAACCGAA |
| 137 | CTGACCAACTTTGAAAGAGGACAGATGAACGGTGTACA |

| | |
|---|---|
| 138 | GACCAGGCGCATAGGCTGGCTGACCTTCATCAAGAGTA |
| 139 | ATCTTGACAAGAACCGGATATTCATTACCCAAATCAAC |
| 140 | GTAACAAAGCTGCTCATTCAGTGAATAAGGCTTGCCCT |
| 141 | GACGAGAAACACCAGAACGAGTAGTAAATTGGGCTTGA |
| 142 | GATGGTTTAATTTCAACTTTAATCATTGTGAATTACCT |
| 143 | TATGCGATTTTAAGAACTGGCTCATTATACCAGTCAGG |
| 144 | ACGTTGGGAAGAAAAATCTACGTTAATAAAACGAACTA |
| 145 | ACGGAACAACATTATTACAGGTAGAAAGATTCATCAGT |
| 146 | TGAGATTTAGGAATACCACATTCAACTAATGCAGATAC |
| 147 | ATAACGCCAAAAGGAATTACGAGGCATAGTAAGAGCAA |
| 148 | CACTATCATAACCCTCGTTTACCAGACGACGATAAAAA |
| 149 | CCAAAATAGCGAGAGGCTTTTGCAAAAGAAGTTTTGCC |
| 150 | AGAGGGGGTAATAGTAAAATGTTTAGACTGGATAGCGT |
| 151 | CCAATACTGCGGAATCGTCATAAATATTCATTGAATCC |
| 152 | CCCTCAAATGCTTTAAACAGTTCAGAAAACGAGAATGA |
| 153 | CCATAAATCAAAAATCAGGTCTTTACCCTGACTATTAT |
| 154 | AGTCAGAAGCAAAGCGGATTGCATCAAAAAGATTAAGA |
| 155 | GGAAGCCCGAAAGACTTCAAATATCGCGTTTTAATTCG |
| 156 | AGCTTCAAAGCGAACCAGACCGGAAGCAAACTCCAACA |
| 157 | GGTCAGGATTAGAGAGTACCTTTAATTGCTCCTTTTGA |
| 158 | TAAGAGGTCATTTTTGCGGATGGCTTAGAGCTTAATTG |
| 159 | CTGAATATAATGCTGTAGCTCAACATGTTTTAAATATG |
| 160 | CAACTAAAGTACGGTGTCTGGAAGTTTCATTCCATATA |
| 161 | ACAGTTGATTCCCAATTCTGCGAACGAGTAGATTTAGT |
| 162 | TTGACCATTAGATACATTTCGCAAATGGTCAATAACCT |
| 163 | GTTTAGCTATATTTTCATTTGGGGCGCGAGCTGAAAAG |
| 164 | GTGGCATCAATTCTACTAATAGTAGTAGCATTAACATC |
| 165 | CAATAAATCATACAGGCAAGGCAAAGAATTAGCAAAAT |
| 166 | TAAGCAATAAAGCCTCAGAGCATAAAGCTAAATCGGTT |
| 167 | GTACCAAAAACATTATGACCCTGTAATACTTTTGCGGG |
| 168 | AGAAGCCTTTATTTCAACGCAAGGATAAAAATTTTTAG |
| 169 | AACCCTCATATATTTTAAATGCAATGCCTGAGTAATGT |
| 170 | GTAGGTAAAGATTCAAAAGGGTGAGAAAGGCCGGAGAC |
| 171 | AGTCAAATCACCATCAATATGATATTCAACCGTTCTAG |
| 172 | CTGATAAATTAATGCCGGAGAGGGTAGCTATTTTTGAG |
| 173 | AGATCTACAAAGGCTATCAGGTCATTGCCTGAGAGTCT |
| 174 | GGAGCAAACAAGAGAATCGATGAACGGTAATCGTAAAA |
| 175 | CTAGCATGTCAATCATATGTACCCCGGTTGATAATCAG |
| 176 | AAAAGCCCCAAAAACAGGAAGATTGTATAAGCAAATAT |
| 177 | TTAAATTGTAAACGTTAATATTTTGTTAAAATTCGCAT |
| 178 | TAAATTTTTGTTAAATCAGCTCATTTTTTAACCAATAG |
| 179 | GAACGCCATCAAAAATAATTCGCGTCTGGCCTTCCTGT |
| 180 | AGCCAGCTTTCATCAACATTAAATGTGAGCGAGTAACA |
| 181 | ACCCGTCGGATTCTCCGTGGGAACAAACGGCGGATTGA |
| 182 | CCGTAATGGGATAGGTCACGTTGGTGTAGATGGGCGCA |
| 183 | TCGTAACCGTGCATCTGCCAGTTTGAGGGGACGACGAC |
| 184 | AGTATCGGCCTCAGGAAGATCGCACTCCAGCCAGCTTT |

| | |
|---|---|
| 185 | CCGGCACCGCTTCTGGTGCCGGAAACCAGGCAAAGCGC |
| 186 | CATTCGCCATTCAGGCTGCGCAACTGTTGGGAAGGGCG |
| 187 | ATCGGTGCGGGCCTCTTCGCTATTACGCCAGCTGGCGA |
| 188 | AAGGGGGATGTGCTGCAAGGCGATTAAGTTGGGTAACG |
| 189 | CCAGGGTTTTCCCAGTCACGACGTTGTAAAACGACGGC |
| 190 | CAGTGCCAAGCTTGCATGCCTGCAGGTCGACTCTAGAGGATCTTTT |

**Supplementary Table S3.** Oligonucleotides were replaced from Table S2 to assemble reference structures. The DNA dumbbell forming sequence is indicated in red.

| Name | Sequence (5' → 3') | Length (nt) | # of oligos replaced |
|---|---|---|---|
| REF 1.1 | ACATCACTTGCCTGAGTAGA | 20 | 26-30 |

| | | | |
|---|---|---|---|
| REF 1.2 | AGAACTCAAATCCTCTTTTGAGGAACAAGTTTTCTTGT CTATCGGCCT | 48 | |
| REF 1.3 | TGCTGGTAATTCCTCTTTTGAGGAACAAGTTTTCTTGT ATCCAGAACA | 48 | |
| REF 1.4 | ATATTACCGCTCCTCTTTTGAGGAACAAGTTTTCTTGT CAGCCATTGC | 48 | |
| REF 1.5 | AACAGGAAAATCCTCTTTTGAGGAACAAGTTTTCTTGT ACGCTCATGG | 48 | |
| REF 1.6 | AAATACCTACTCCTCTTTTGAGGAACAAGTTTTCTTGT ATTTTGACGC | 48 | |
| REF 1.7 | TCAATCGTCTTCCTCTTTTGAGGAACAAGTTTTCTTGT GAAATGGATT | 48 | |
| REF 1.8 | ATTTACATTGGCAGATTCAC | 20 | |
| REF 1.9 | CAGTCACACGACCAGTAATAAAAGGGACAT | 30 | |
| REF 2.1 | TCACAAACAAATAAATCCTCATTAAAGCCAGAATGGAA AGCGCAGTCTCTGAATTT | 56 | |
| REF 2.2 | ACCGTTCCAGTAAGCGTCAT | 20 | |
| REF 2.3 | ACATGGCTTTTCCTCTTTTGAGGAACAAGTTTTCTTGT TGATGATACA | 48 | |
| REF 2.4 | GGAGTGTACTTCCTCTTTTGAGGAACAAGTTTTCTTGT GGTAATAAGT | 48 | |
| REF 2.5 | TTTAACGGGGTCCTCTTTTGAGGAACAAGTTTTCTTGT TCAGTGCCTT | 48 | 106-112 |
| REF 2.6 | GAGTAACAGTTCCTCTTTTGAGGAACAAGTTTTCTTGT GCCCGTATAA | 48 | |
| REF 2.7 | ACAGTTAATGTCCTCTTTTGAGGAACAAGTTTTCTTGT CCCCCTGCCT | 48 | |
| REF 2.8 | ATTTCGGAACTCCTCTTTTGAGGAACAAGTTTTCTTGT CTATTATTCT | 48 | |
| REF 2.9 | GAAACATGAAAGTATTAAGA | 20 | |
| REF 2.10 | GGCTGAGACTCCTCAAGAGAAGGATTAGGATTAGCGGG GTTTTGCTCAGT | 50 | |

### Section S4. AFM imaging

#### *Section 4.1 Sample preparation and imaging with AFM*

Atomic Force Microscopy (Nanosurf Mobile S) imaging of nanobaits was performed in the air in a non-contact mode. All scans were performed on a bare mica surface following the adsorption of nanobaits as follows. We placed a 10 μL drop of a DNA solution (diluted to 1 ng/μL in filtered 10 mM $MgCl_2$ and 10 mM Tris-HCl, pH 8.0) onto a freshly cleaved mica surface for 1 minute, rinsed the mica plate three times with 100 μL of Mili-Q water and then blow-dried it with Nitrogen. Before the scan, the mica plate was affixed to the AFM sample stage using double-sided adhesive tape. Image visualization and analysis were done using Gwyddion.

#### *Section 4.2 AFM images of nanobait*

AFM images of nanobait are shown in Figures S3 and S4. Figure S3a presents AFM images of the nanobait structure without any label added. In some cases, two references are visible (circled in dark grey, Figure S3a, right). However, it is known that DNA dumbbell structures are hardly distinguishable with AFM imaging in comparison to nanopores [1].

Nanobait events with monovalent streptavidin (Figure S3b) indicate five structures that correspond to capture sites with labels, as expected from the nanobait design. The same design but with the DNA flower label is presented in Figure S4. The DNA flower is larger than streptavidin and hence easier to be detected using AFM imaging (Figure S2).

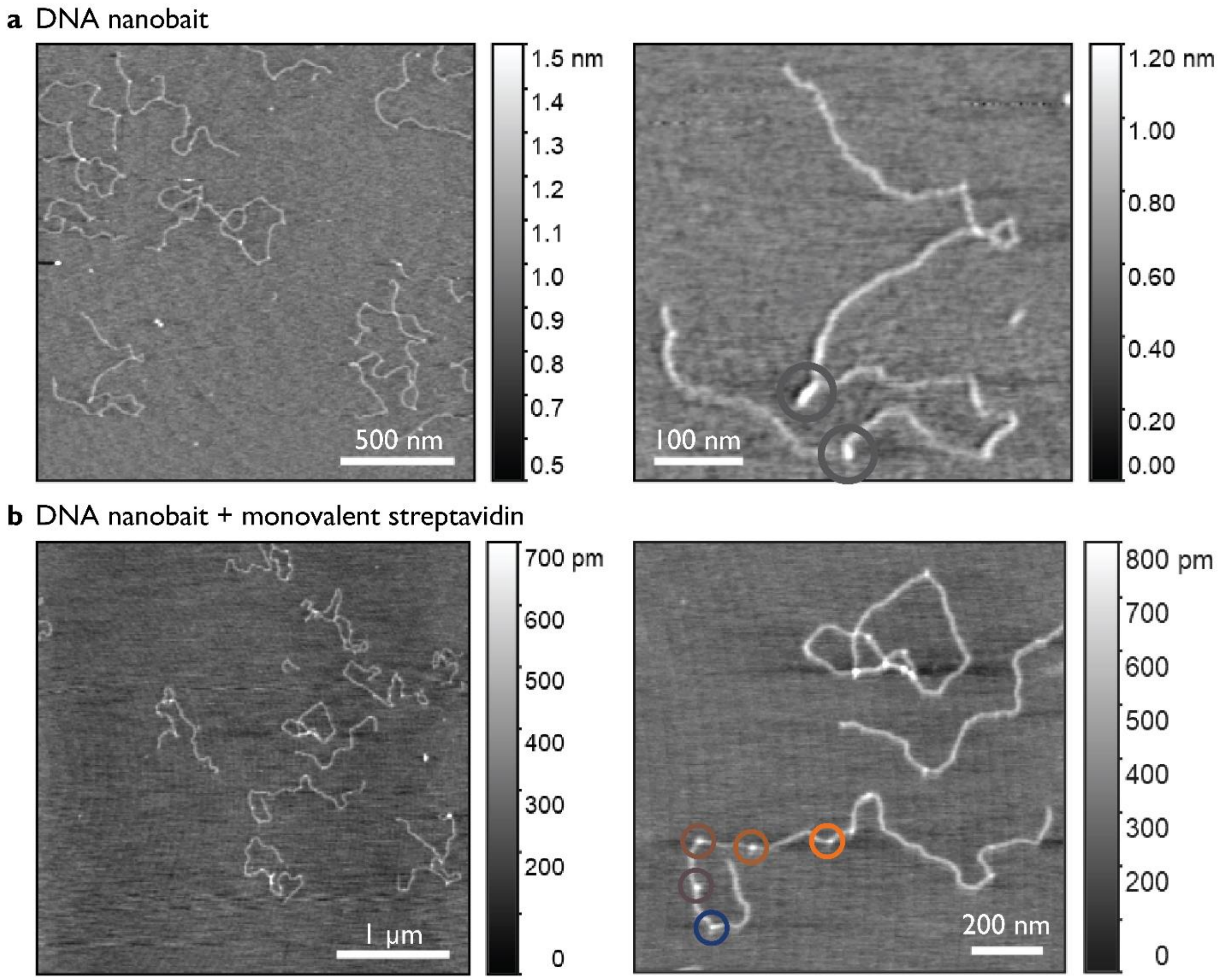

**Figure S3.** AFM images of DNA nanobait (as in Figure 3) with monovalent streptavidin. a) Nanobait without monovalent streptavidin has two reference structures (circled in dark gray). b) AFM images of nanobait with monovalent streptavidin used as a label. Five capture sites are circled in corresponding colors. Five non-overlapping areas are imaged for each sample.

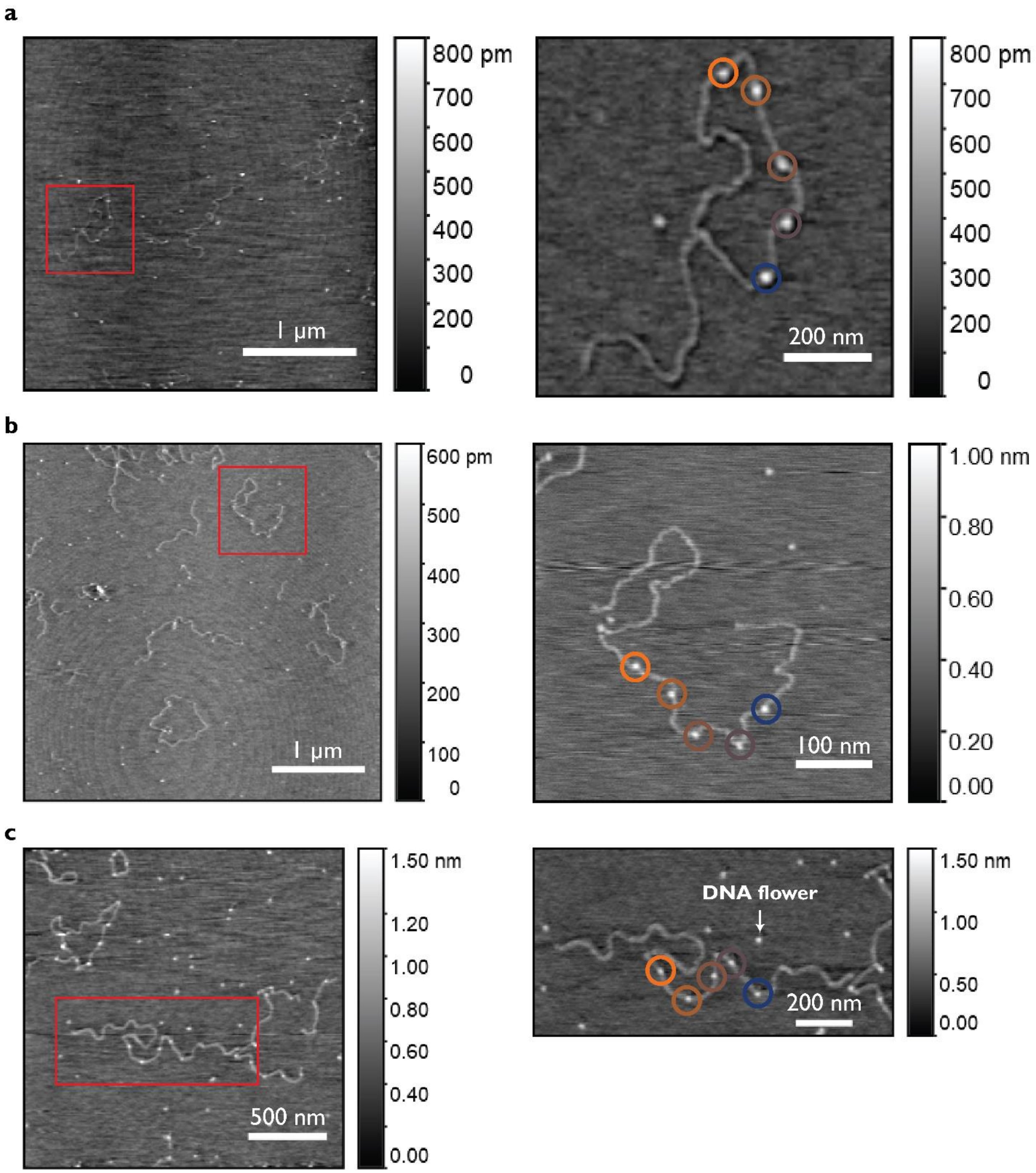

**Figure S4.** AFM images of nanobait (as in Figure 3) with DNA flower as a label. Three different nanobait example images are shown in a), b), and c). Five non-overlapping areas are imaged for each sample.

### Section S5. EMSA analysis

We performed the electrophoretic mobility shift assay (EMSA) to see a mobility shift caused by streptavidin binding to all five sites on nanobait (Figure S5). 2 % (w/v) agarose (BioReagent, low electroendosmosis, Merck SigmaAldrich; catalog number A9539) gel was prepared by adding 2 g of agarose, 10 mL of 10 × Tris-Borate-EDTA buffer (TBE buffer), and Milli-Q water was added to 100 mL. The gel was heated in the microwave at the maximum power (800 W) for 2-3 min. It was cooled down and poured into a gel tray to set for 45 min.

Samples were mixed with purple gel loading dye (New England Biolabs; catalog number B7025S) and TBE buffer to 1 × with ~150 ng of a DNA sample. We used a 1 kb DNA ladder as a reference (New England Biolabs; catalog number N3232) with a size range from 500 bp to 10 kb (Figure S5a). Nanobait in lane 2 (Figure S5a) was mixed with 10 × excess of wild-type tetravalent streptavidin (ThermoFisher Scientific; catalog number 21125).

It can be observed from the agarose gel in Figure S5a that there is a slight shift from nanobait (lane 1) to nanobait + streptavidin (lane 2). We used gel intensity analysis using an open-source software Fiji[8] to plot this shift, indicating that streptavidin(s) is bound to nanobait (Figure S5b).

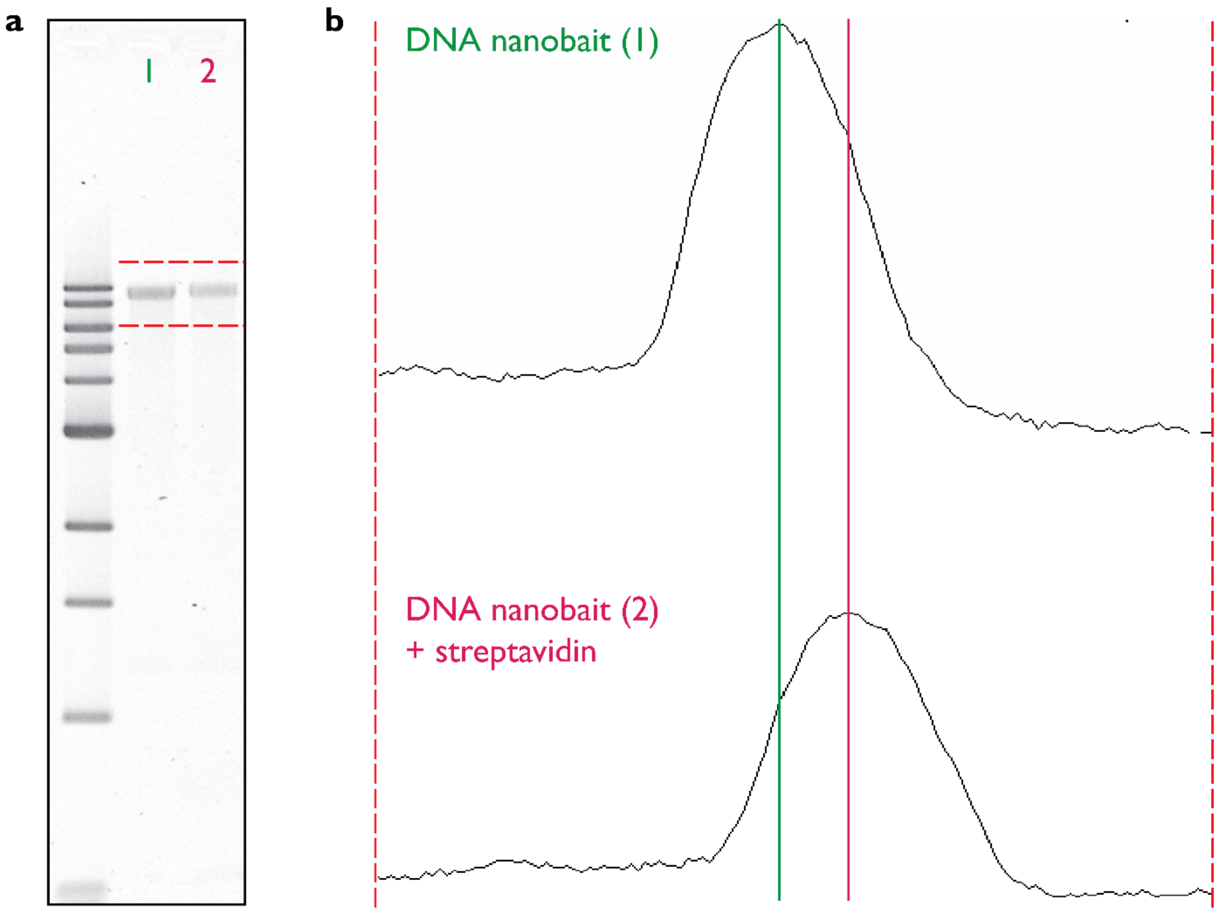

**Figure S5.** Electrophoretic mobility shift assay (EMSA) of nanobait without and with streptavidin. a) 2% (w/v) agarose gel od nanobait without (lane 1) and with (lane 2) streptavidin added. We used a 1 kb DNA ladder (New England Biolabs; product number N3232). The two top lines of the ladder are 10 and 8 kb. b) The intensity traces of corresponding lanes from a). The green line indicates the intensity maximum of nanobait (lane 1) and the purple line indicates the intensity maximum of nanobait + streptavidin (lane 2). The shift between the green and purple lines corresponds to streptavidin(s) bound to capture sites on nanobait. Samples were run once.

### Section S6. Multiple respiratory viruses DNA nanobait sequences and example events

Nanobait for multiple respiratory viruses is prepared as previously described in Section S2. Below, are listed sequences of capture sites (Table S7), biotin strand (Table S6), and target sequence (Table S5) for each virus.

We also show here additional nanopore events of nanobait without target added (Figure S6a), with SARS-CoV-2 target (Figure S6b), Influenza A (Figure S6c), RSV (Figure S6d), Parainfluenza (Figure S6e), and Rhinoviruses (Figure S6f). Targets for Influenza A, Parainfluenza A, and Rhinoviruses target cohort rather than a single variant [9,10]. The nanobait and peaks resemble those presented in Figure 2a of the manuscript. These single-molecule events are obtained from three separate nanopore measurements.

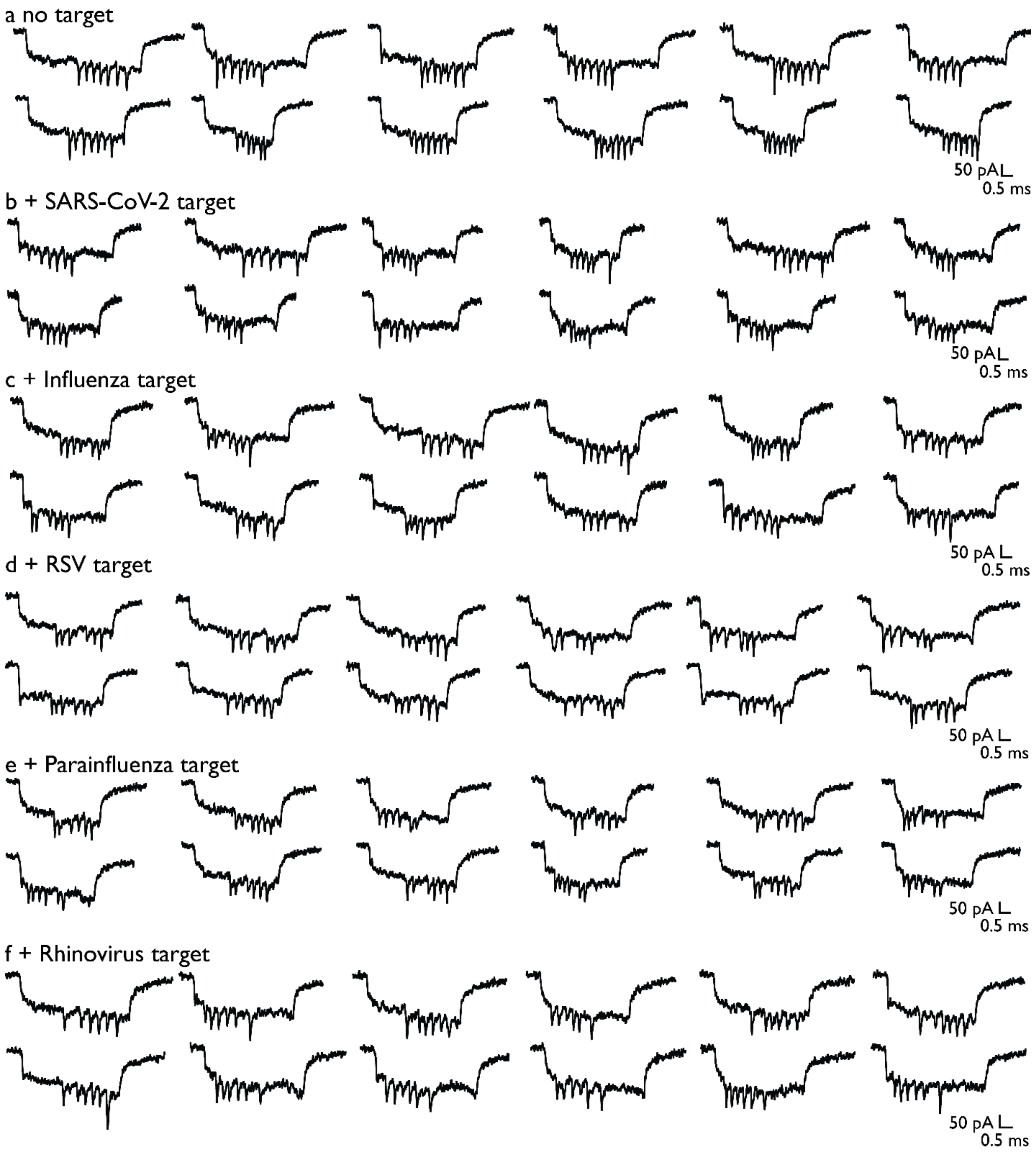

**Figure S6.** Additional example events for nanobait used for identification of various respiratory viruses with a) no target added, b) SARS-CoV-2 target, c) Influenza target, d) RSV, e) Parainfluenza target, and f) Rhinovirus target.

**Supplementary Table S4.** Presence of peaks at their respective sites for multiple virus identification.

| Target/Case | Control (no targets) | Standard error | Target present | Standard error |
|---|---|---|---|---|
| SARS-CoV-2 | 0.17 | 0.04 | 0.75 | 0.10 |
| RSV | 0.09 | 0.01 | 0.96 | 0.06 |
| Rhinovirus | 0.08 | 0.03 | 0.89 | 0.13 |
| Influenza | 0.11 | 0.04 | 0.49 | 0.17 |
| Parainfluenza | 0.03 | 0.01 | 0.73 | 0.04 |

**Supplementary Table S5.** Target sequences for multiple virus identification.

| Strand name | Virus / group of viruses | Sequence reference | Sequence (5' → 3') |
|---|---|---|---|
| SW | SARS-CoV-2_Reference | NCBI Reference Sequence: NC_045512.2[11] | GTATGAAAATGCCTTTTTAC |
| Infl | Influenza A viruses universal | Sequence adapted from[12] | TGACAGGATTGGTCTTGTCT |
| RSV | Respiratory syncytial virus universal A | Sequence adapted from[13] | ACACAGCAGCTGTTCAGTAC |
| PI | Parainfluenza 1 | Sequence adapted from[10] | CTTCCTGCTGGTGTGTTAAT |
| RV | Rhinoviruses universal | Sequence adapted from[9] | TCCTCCGGCCCCTGAATGTG |

**Supplementary Table S6.** 3' biotinylated strand sequences for multiple virus identification.

| Strand name | Virus / group of viruses | Sequence (5' → 3') |
|---|---|---|
| bSW | SARS-CoV-2_Reference | AAATGCCTTTTTAC/3-biotin/ |
| bInfl | Influenza A viruses universal | GATTGGTCTTGTCT/3-biotin/ |
| bRSV | Respiratory syncytial virus universal A | CAGCTGTTCAGTAC/3-biotin/ |
| bPI | Parainfluenza 1 | GCTGGTGTGTTAAT/3-biotin/ |

| | | |
|---|---|---|
| bRV | Rhinoviruses universal | GGCCCCTGAATGTG /3-biotin/ |

**Supplementary Table S7.** Capture strand sequences for multiple virus identification.

| Strand name | Virus / group of viruses | Sequence (5’ → 3’) |
|---|---|---|
| cSW_42 | SARS-CoV-2_Reference | TTCGACAACTCGTATTAAATCCTTTGCCCGAACGTTAT TTTTT GTAAAAAGGCATTTTCATAC |
| cRSV_55 | Respiratory syncytial virus universal A | GTGAGTGAATAACCTTGCTTCTGTAAATCGTCGCTATT TTTTT GTACTGAACAGCTGCTGTGT |
| cRV_68 | Rhinoviruses universal | AGAATATAAAGTACCGACAAAAGGTAAAGTAATTCTGT TTTTT CACATTCAGGGGCCGGAGGA |
| cI_81 | Influenza A viruses universal | TCCCAATCCAAATAAGAAACGATTTTTTGTTTAACGTC TTTTT AGACAAGACCAATCCTGTCA |
| cPI_94 | Parainfluenza 1 | CATTCAACCGATTGAGGGAGGGAAGGTAAATATTGACG TTTTT ATTAACACACCAGCAGGAAG |

**Section S7. Multiple SARS-Co-V-2 virus variants DNA nanobait sequences and example events**

Nanobait for multiple SARS-CoV-2 variants was prepared as previously described in Section S2. Below are listed sequences of capture sites (Table S12), biotin strand (Table S10), and target sequence of the wildtype (Table S11) and variant (Table S9).

We also showed here additional nanopore events of nanobait without target added (Figure S7a), with B reference target (Figure S7b), B.1.617 variant target (Figure S7c), B.1 variant target (Figure S7d), B.1.1.7 variant target (Figure S7e), and B.1.351 variant target (Figure S7f). The nanobait and peaks resembled those presented in Figure 2b of the manuscript. These single-molecule events were obtained from three separate nanopore measurements.

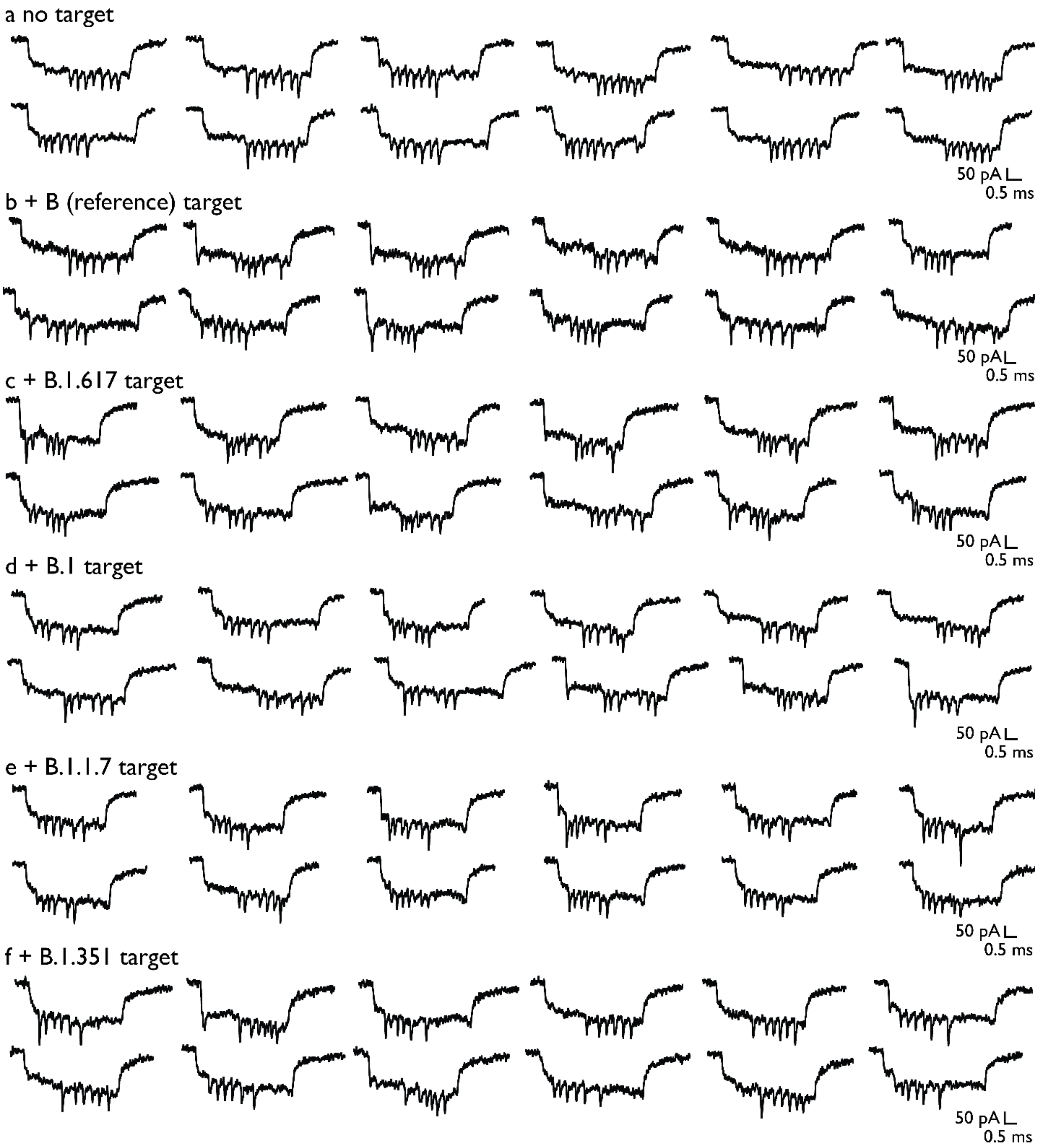

**Figure S7.** Additional example events for nanobait used for identification of various SARS-CoV-2 variants with a) no target added, b) B target, c) B.1.617 variant target, d) B.1 variant target, e) B.1.1.7 variant target, and f) B.1.351 variant target.

**Supplementary Table S8.** Presence of peaks at their respective sites for multiple variant identification.

| Target/Case | Control (no targets) | Standard error | WT target present | Standard error | Variant target present | Standard error |
|---|---|---|---|---|---|---|
| B (reference) | 0.08 | 0.00 | 0.93 | 0.04 | 0.92 | 0.06 |
| B.1.617 (Delta) | 0.23 | 0.01 | 0.39 | 0.02 | 0.77 | 0.01 |
| B.1 | 0.02 | 0.00 | 0.29 | 0.05 | 0.98 | 0.06 |
| B.1.1.7 (Alpha) | 0.04 | 0.00 | 0.06 | 0.02 | 0.96 | 0.12 |
| B.1.351 (Beta) | 0.10 | 0.00 | 0.49 | 0.11 | 0.90 | 0.03 |

**Supplementary Table S9.** Target sequences for multiple SARS-CoV-2 variant identification. In red, single nucleotide variant positions are highlighted, while yellow highlights the toehold region used for the SDR.

| Strand name | WHO nomenclature[14] | Pangolin nomenclature[15] | Single nucleotide variation | Sequence (5' → 3') |
|---|---|---|---|---|
| SW | Reference | B (reference) | / | GTATGAAAATGCCTTTTTAC |
| SIm | Delta | B.1.617 | T-G<br>L452R | CCGGTATAGATTGTTTAGGA |
| SEm | NA | B.1 | A-G<br>D614G | GGTGTTAACTGCACAGAAGT |
| SUKm | Alpha | B.1.1.7 | A-T<br>N501Y | CTTATGGTGTTGGTTACCAA |
| SSAm | Beta | B.1.351 | G-A<br>E484K | TAAAGGTTTTAATTGTTACT |

**Supplementary Table S10.** 3' biotinylated strand sequences for multiple SARS-CoV-2 variant identification.

| Strand name | Variant name | Sequence (5’ → 3’) |
|---|---|---|
| bSW | B (reference) | AAATGCCTTTTTAC/3-BIOTIN/ |
| bSIm | B.1.617 (Delta) | TAGATTGTTTAGGA/3-BIOTIN/ |
| bSEm | B.1 | AACTGCACAGAAGT/3-BIOTIN/ |
| bSUKm | B.1.1.7 (Alpha) | GTGTTGGTTACCAA/3-BIOTIN/ |
| bSSAm | B.1.351 (Beta) | TTTTAATTGTTACT/3-BIOTIN/ |

**Supplementary Table S11.** Wild-type (WT) target sequences for multiple SARS-CoV-2 variant identification. In green, wild-type nucleotide positions are highlighted, while yellow highlights the toehold region used for the SDR.

| Strand name | Sequence (5’ → 3’) | Length (nt) |
|---|---|---|
| SI | CCTGTATAGATTGTTTAGGA | 20 |
| SE | GATGTTAACTGCACAGAAGT | 20 |
| SUK | CTAATGGTGTTGGTTACCAA | 20 |
| SSA | TGAAGGTTTTAATTGTTACT | 20 |

**Supplementary Table S12.** Capture strand sequences for multiple SARS-CoV-2 variant identification.

| Strand name | Variant name | Sequence (5’ → 3’) |
|---|---|---|
| cSW_42 | B (reference) | TTCGACAACTCGTATTAAATCCTTTGCCCGAACGTTAT TTTTT GTAAAAAGGCATTTTCATAC |
| cSI_68 | B.1.617 (Delta) | AGAATATAAAGTACCGACAAAAGGTAAAGTAATTCTGT TTTTT TCCTAAACAATCTATACCGG |
| cSE_94 | B.1 | CATTCAACCGATTGAGGGAGGGAAGGTAAATATTGACG TTTTT ACTTCTGTGCAGTTAACACC |
| cSUK_81 | B.1.1.7 (Alpha) | TCCCAATCCAAATAAGAAACGATTTTTTGTTTAACGTC TTTTT TTGGTAACCAACACCATAAG |

| | | |
|---|---|---|
| cSSA_55 | B.1.351 (Beta) | GTGAGTGAATAACCTTGCTTCTGTAAATCGTCGCTATT TTTTT AGTAACAATTAAAACCTTTA |

#### Section S8. Discrimination of control SARS-Co-V-2 RNA virus variants

Nanobait for SARS-CoV-2 N501 RNA virus variants was prepared as previously described in Section S2 (Figure S8). Below, are listed sequences of SARS-CoV-2 N501 RNA, capture sites, biotinylated strand, and guide oligos used for RNase H cutting (Table S13).

*Programmable RNase H cutting of SARS-CoV-2 RNA controls for nanobait*

For nanopore sensing, SARS-CoV-2 RNA (S:N501 in Table S13) controls were used for the detection with nanobait. Firstly, we mixed guide oligos with a SARS-CoV-2 N501 RNA in the ratio 1:1:1 and heated the mixture to 70 °C for 5 minutes. RNase H (5,000 units/ml, NEB) was added, mixed, and heated for 20 minutes at 37 °C to allow the enzyme to cut RNA in the DNA : RNA hybrid that effectively leads to the release of target RNA. RNase H is thermally inactivated by incubation at 65 °C for 10 min.

*Nanopore readout of DNA nanobait*

Nanobait was mixed with a cut long RNA control at ten times excess in 10 mM $MgCl_2$ and 100 mM NaCl. The mixture (5 μL) was incubated at room temperature (~10 min) until prepared for nanopore measurement.

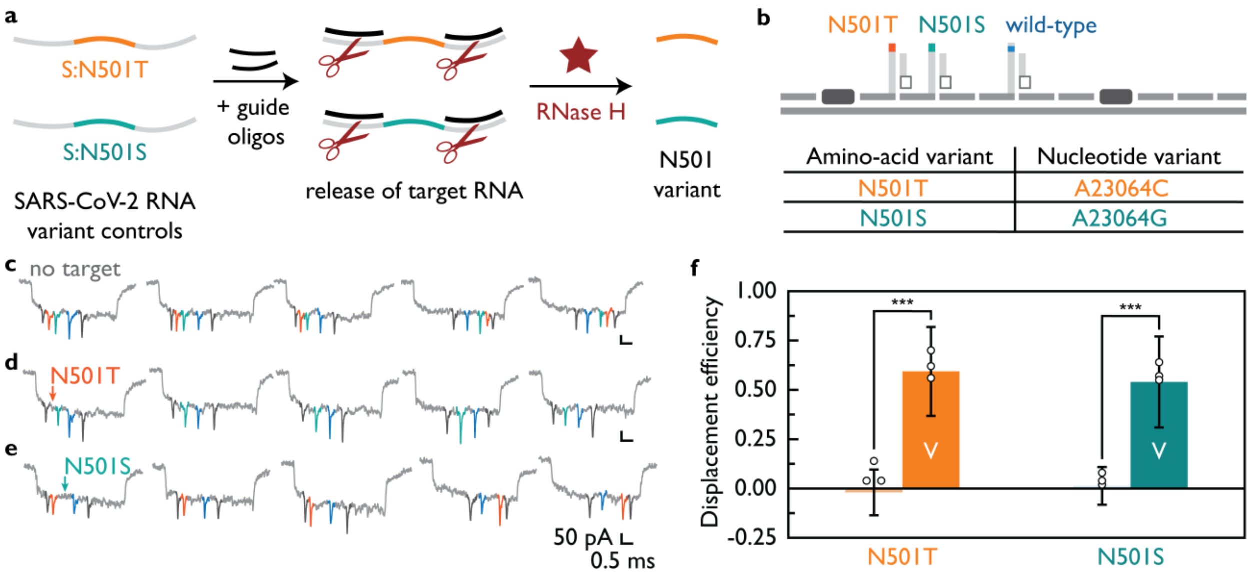

**Figure S8. Nanobait discriminates single-nucleotide SARS-CoV-2 RNA variants.** a) SARS-CoV-2 RNA variant controls that have one amino-acid substitutions. Guide oligos are added next to the target RNA sequence hence enabling RNase H cutting and releasing of target RNAs in solution. b) Nanobait design has two references, two sites for each of variant (N501T, N501S) and a wild-type site. In the table each of these amino-acid variants are shown with position and substitution of nucleotide counterpart indicated. c)

Example events for no target control indicate the correct number of downward spikes each corresponding to a structure depicted in b). In d) and e) are shown example events for each of the N501 variants. The absence of the colored spike indicates the presence of each respective target. f) Displacement efficiencies for single-nucleotide SARS-CoV-2 variants (labelled as 'V') are compared with the displacement efficiency for the wild-type SARS-CoV-2 (B.1.1.7). Error bars represent standard error and the center as the mean for three nanopore measurements and fifty nanopore events per measurement. The difference between conditions without and with variant targets is statistically significant (***$p < 0.001$; two-sided Student's T test; N=150).

**Supplementary Table S13.** SARS-CoV-2 RNA sequences and guide oligos, capture sites and biotinylated strands.

| Strand name | Description | Sequence (5' → 3') |
|---|---|---|
| S:N501_RNA | Wild-type oligo | ATCATATGGTTTCCAACCCA CTTATGGTGTTGGTTACCAA CCATACAGAGTAGTAGTACT |
| S:N501T_RNA | A23064C | ATCATATGGTTTCCAACCCA CTTCTGGTGTTGGTTACCAA CCATACAGAGTAGTAGTACT |
| S:N501S_RNA | A23064G | ATCATATGGTTTCCAACCCA CTTGTGGTGTTGGTTACCAA CCATACAGAGTAGTAGTACT |
| Guide oligo S:N501_a | Downstream cutting oligo | TGGGTTGGAAACCATATGAT |
| Guide oligo S:N501_b | Upstream cutting oligo | AGTACTACTACTCTGTATGG |
| cS42:N501T | Carrier overhang | TTCGACAACTCGTATTAAATCCTTTGCCCGAACGTTAT TTTTT |
| cS55:N501S | Carrier overhang | GTGAGTGAATAACCTTGCTTCTGTAAATCGTCGCTATT TTTTT |
| cSUK_81 | Carrier overhang | TCCCAATCCAAATAAGAAACGATTTTTTGTTTAACGTC TTTTT TTGGTAACCAACACCAAAAG |
| bS:N501 | Biotin | GTGTTGGTTACCAA/3-BIOTIN/ |

### Section S9. Kinetics of SDR with DNA and RNA targets

Nanobait for multiple SARS-CoV-2 targets is prepared as previously described in Section S2. Below, are listed sequences of capture sites (Table S17), biotin strands (Table S16), and target strands (Table S15).

Kinetics measurements for all five targets SDR are shown in Figure S9 and fifty nanobait events are analyzed per each data point. We tested targets of the same sequence but with different backbones as RNAs or DNAs (Figure S9a and S9b, respectively). Here, we show that target chemistry might have an effect on the kinetics of the SDR since some of the targets (e.g. target 3) have slower SDR for the RNA sequence than compared to the DNA sequence.

Additional nanopore events of nanobait without any target added (Figure S9c) and all five targets present (Figure S9d).

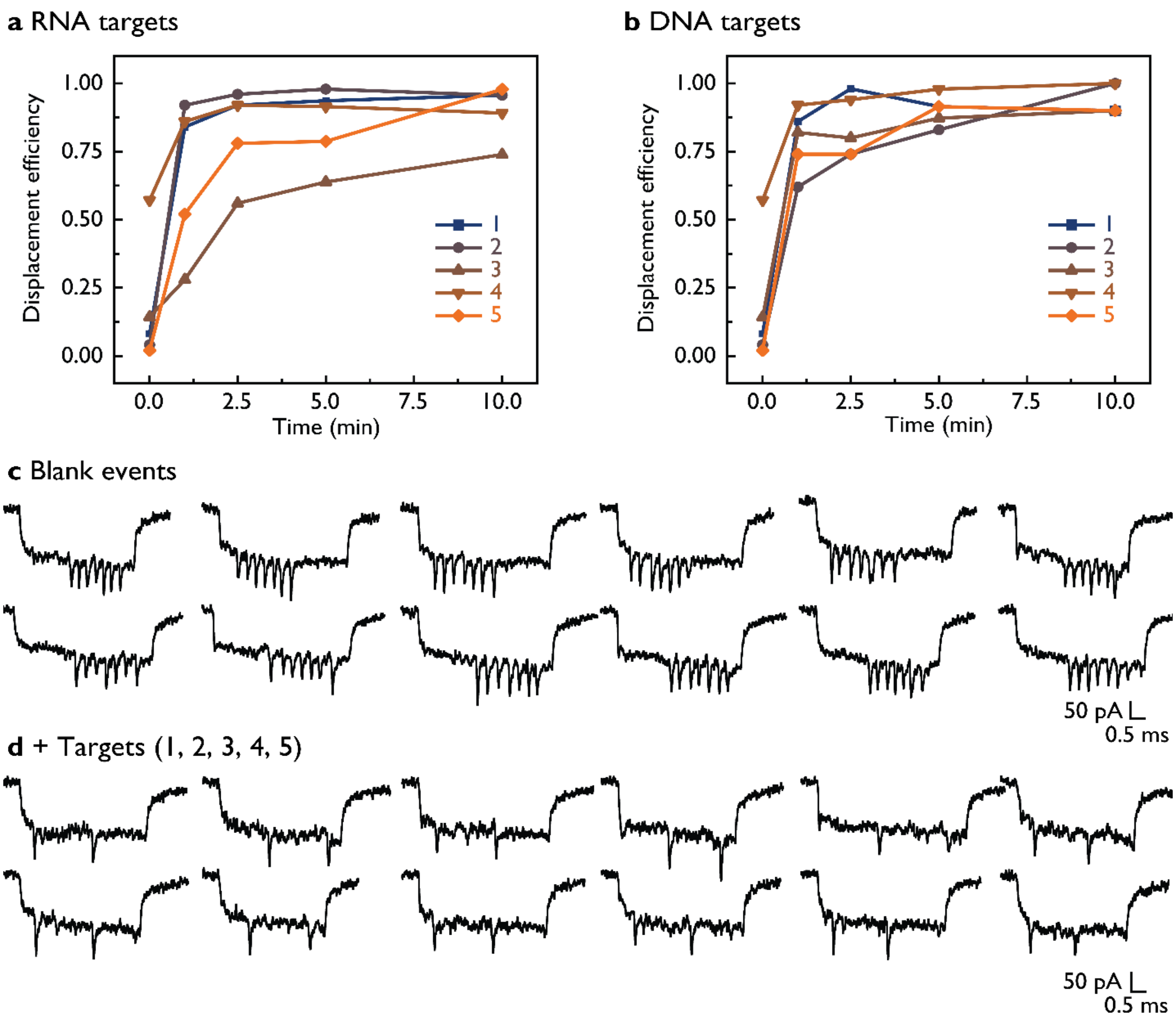

**Figure S9.** Kinetics of the toehold strand-displacement reaction (SDR) using RNA and DNA targets. Displacement efficiency for five different RNA or DNA targets is plotted in a) and b), respectively. c) Blank (no targets added) nanopore events indicate correct nanobait assembly. d) After the SDR with five targets added all five target-specific downward signals diminish.

We have measured single-molecule kinetics of SDR over 9 h for RNA targets (Figure S10). This experiment is performed with the diluted nanobait sample and in 1:1 ratio nanobait to target and allowed the typical 10 min incubation for the SDR. We have obtained from 1150 to 2860 events per hour slot (~18000 events) and the first fifty events were analyzed after each hour and the average value is plotted in Figure S10.

We can observe a general trend of increasing displacement efficiency for all five targets (Figure S10).

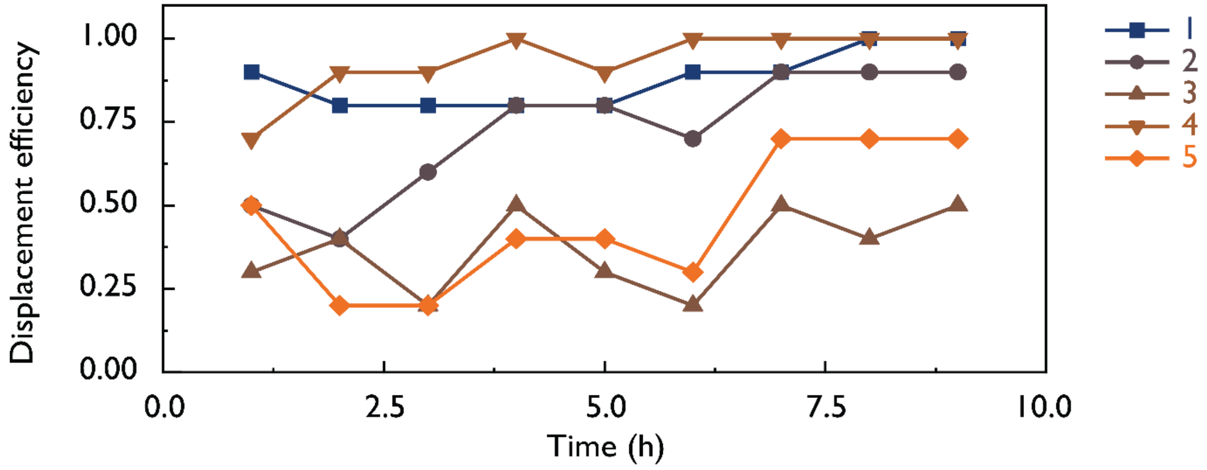

**Figure S10.** Single-molecule kinetics of the toehold strand-displacement reaction (SDR) using RNA targets.

**Supplementary Table S14.** Presence of peaks at their respective sites for multiple SARS-CoV-2 target identification.

| Target/Case | Control (no targets) | Standard error | Target present | Standard error |
|---|---|---|---|---|
| H1 | 0.08 | 0.14 | 0.92 | 0.08 |
| H2 | 0.10 | 0.16 | 0.90 | 0.22 |
| H3 | 0.13 | 0.18 | 0.87 | 0.14 |
| H4 | 0.38 | 0.11 | 0.62 | 0.08 |
| H5 | 0.08 | 0.14 | 0.92 | 0.12 |

**Supplementary Table S15.** Target sequences for multiple SARS-CoV-2 target identification.

| Strand name | Sequence (5' → 3') | Length (nt) |
|---|---|---|

| | | |
|---|---|---|
| H1 | TGATTGTGAAGAAGAAGAGT | 20 |
| H2 | AAGAAAGGAGCTAAATTGTT | 20 |
| H3 | AGAGTTGATTTTTGTGGAAA | 20 |
| H4 | TGGTGTTTATTCTGTTATTT | 20 |
| H5 | GGTAAAGTTGAGGGTTGTAT | 20 |

**Supplementary Table S16.** 3’ Biotinylated strand sequences for SARS-CoV-2 target identification in a patient sample.

| Strand name | Sequence (5’ → 3’) | Length (nt) |
|---|---|---|
| bH1 | TGAAGAAGAAGAGT /3’-biotin/ | 14 |
| bH2 | GGAGCTAAATTGTT /3’-biotin/ | 14 |
| bH3 | GATTTTTGTGGAAA /3’-biotin/ | 14 |
| bH4 | TTATTCTGTTATTT /3’-biotin/ | 14 |
| bH5 | GTTGAGGGTTGTAT /3’-biotin/ | 14 |

**Supplementary Table S17.** Capture strand sequences for the multiple SARS-CoV-2 target identification.

| Strand name | Sequence (5’ → 3’) | Length (nt) |
|---|---|---|
| cH1_42 | TTCGACAACTCGTATTAAATCCTTTGCCCGAACGTTAT TTTTT ACTCTTCTTCTTCACAATCA | 63 |
| cH2_55 | GTGAGTGAATAACCTTGCTTCTGTAAATCGTCGCTATT TTTTT AACAATTTAGCTCCTTTCTT | 63 |
| cH3_68 | AGAATATAAAGTACCGACAAAAGGTAAAGTAATTCTGT TTTTT TTTCCACAAAAATCAACTCT | 63 |

| | | |
|---|---|---|
| cH4_81 | TCCCAATCCAAATAAGAAACGATTTTTTGTTTAACGTC TTTTT AAATAACAGAATAAACACCA | 63 |
| cH5_94 | CATTCAACCGATTGAGGGAGGGAAGGTAAATATTGACG TTTTT ATACAACCCTCAACTTTACC | 63 |

**Section S10. Gel analysis of MS2 RNA cutting**

Oligonucleotides A and B (Figure 4c) correspond to guide oligos TXA and TXB (X being the target name). Oligo C corresponds to cTX (X being the target name).

We treated MS2 RNA with a single target or with all three targets together to visualize site-specific cutting of RNA. We analyzed these samples using agarose gel electrophoresis (Figure S11). After MS2 RNA cutting for all three sites, we can observe expected lane mobility.

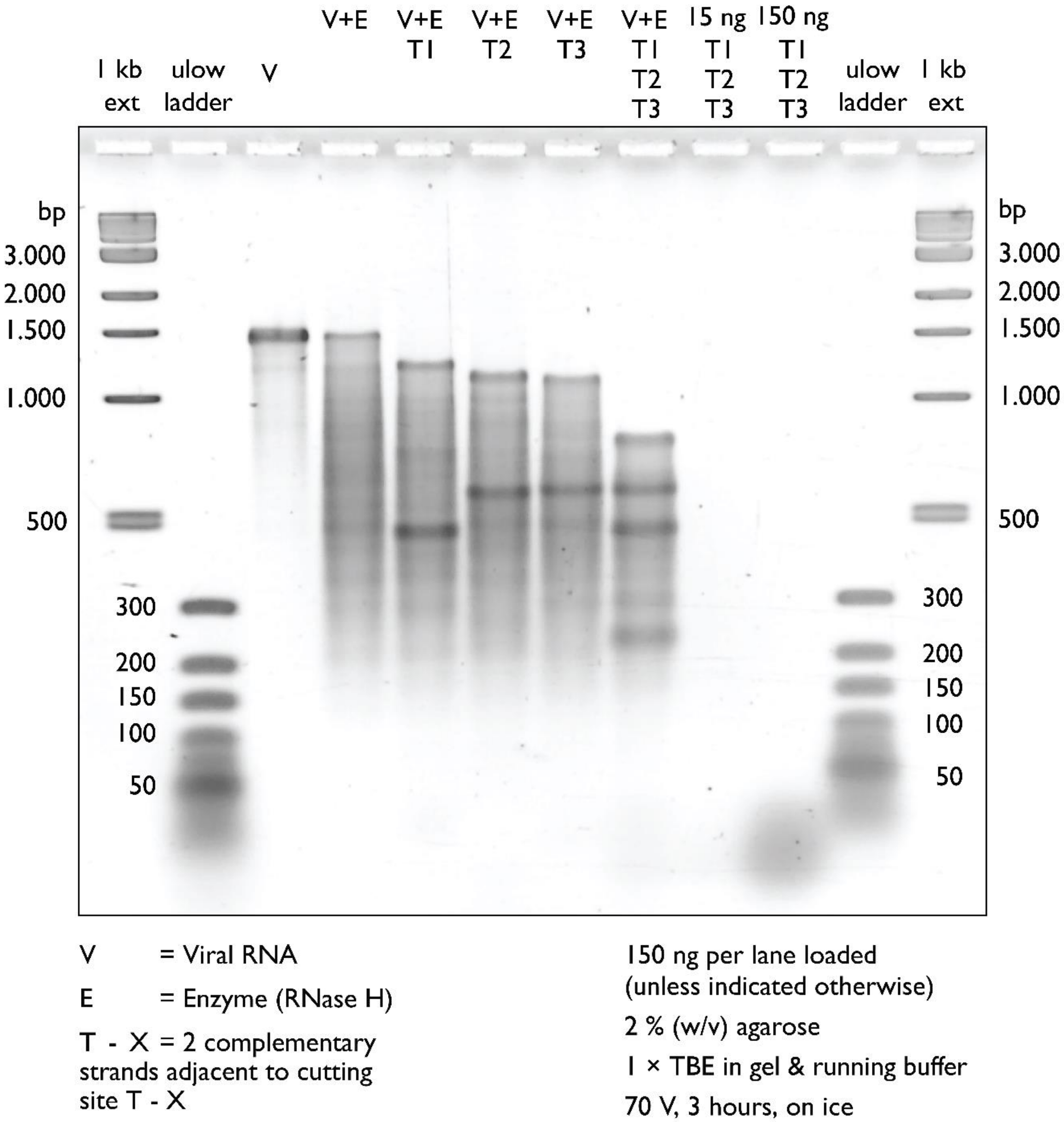

**Figure S11.** Agarose gel analysis of RNase H cutting of MS2 viral RNA.

We tested the mobility of targets, guide oligos, and capture oligos with non-denaturing polyacrylamide gel electrophoresis (PAGE) analysis (Figure S12). Despite targets and guide oligos having the same length they have different band mobility. This result indicates that mobility can be related to the sequence, given that this is the sole differentiating factor. We employed this advantage to detect each target in the background of guide oligos. Samples were run once.

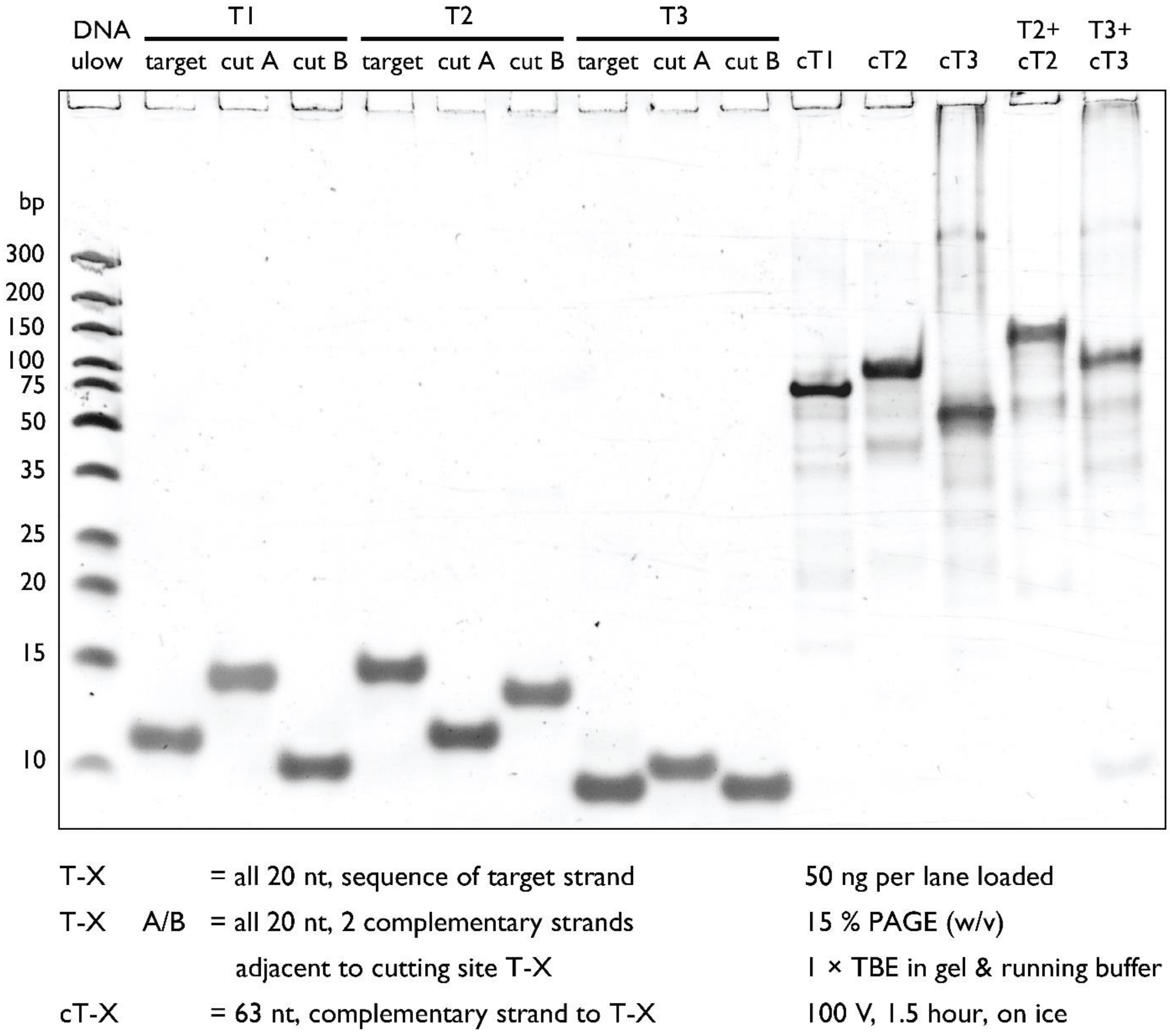

**Figure S12.** PAGE analysis of oligonucleotides used for MS2 RNase H cutting, target strands, and complementary capture strands. Thermo Scientific™ GeneRuler Ultra Low Range DNA Ladder was used (DNA ulow). Samples were run once.

We performed a control PAGE analysis by treating MS2 RNA with RNase H cutting without guide oligos. Here, we tested if targets have significant binding to enzyme-treated RNA (Figure S13).

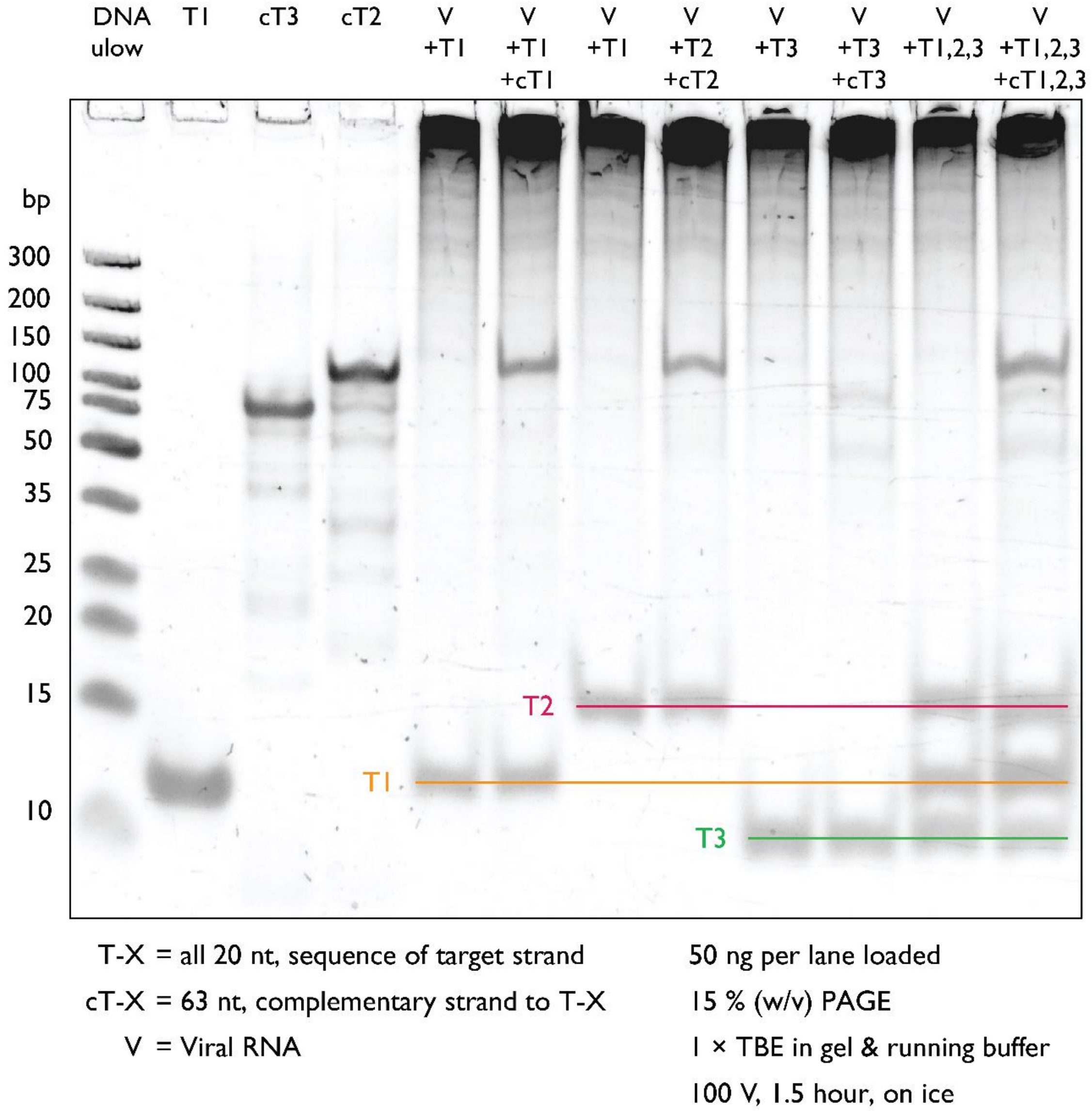

**Figure S13.** PAGE analysis of oligonucleotide mobility in a background of MS2 RNA treated with RNase H without added guide oligos. Samples were run once.

Previously, we have shown that MS2 RNA is cut in fragments of the desired size with 100 % efficiency. However, the release of target RNAs from cut MS2 RNA cannot be tested using this gel. In Figure S14 we tested if RNA target is free in solution. Once MS2 RNA is cut with one guide oligo, the target is absent (x T1A) and the T1A oligo is the only one visible. If both guide oligos are added for target T1 an additional band is visible on the gel. We wanted to validate that this band is the T1 target. This is performed by mixing cut MS2 with a complementary strand (cT1+30 T; the sequence is listed in Table S23) and observing a shift from the cT1+30 T strand.

In the case of the T2 target, we have not observed any additional band indicating that target M2 is probably bound to its position in MS2 RNA structure, as discussed in the main text.

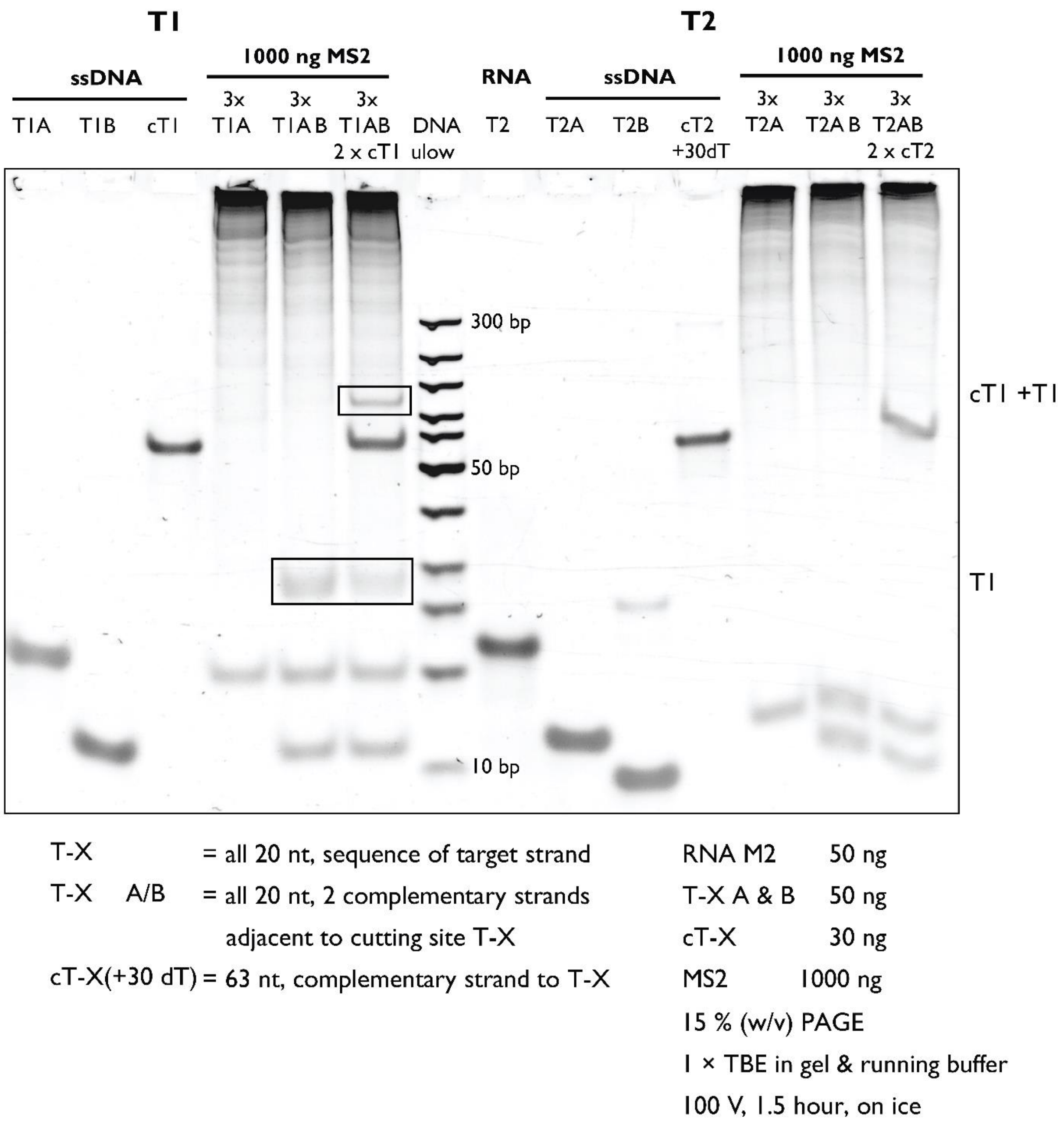

**Figure S14.** PAGE analysis of target detection after MS2 RNA is treated with RNase H with respective guide oligos for targets T1 and T2 added. DNA ulow lane corresponds to GeneRuler Uultra Low Range DNA ladder (Thermo Fisher Scientific) with the lowest band being 10 bp and the highest band being 300 bp in length. Samples were run once.

**Section S11. DNA nanobait for MS2 RNA target detection**

Nanobait for MS2 virus is prepared as previously described in Section S2. Below are listed sequences of capture sites (Table S21), biotin strand (Table S20), and target sequence (Table S19). In Table S22 are listed guide oligos for all targets.

We also show additional nanopore events of nanobait without cut MS2 (Figure S15a), and with cut MS2 RNA added (Figure S15b). The nanobait and peaks resemble those presented in Figure 4b of the manuscript. These single-molecule events are obtained from three separate nanopore measurements.

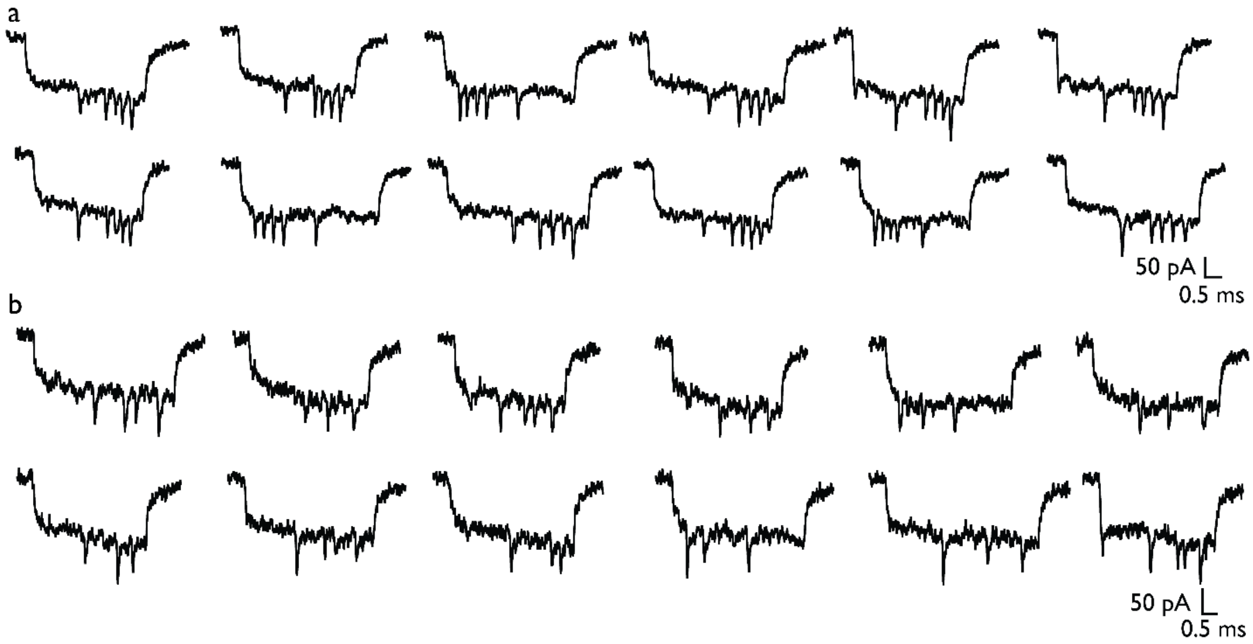

**Figure S15.** Example events for nanobait with cut MS2 RNA. a) Single-molecule nanobait events indicate that a) all three peaks are present, hence targets are not present, or b) peaks are absent, hence targets are present.

**Supplementary Table S18.** Presence of peaks at their respective sites for multiple MS2 viral target identification.

| Target/Case | Control (no targets) | Standard error | Target present | Standard error |
|---|---|---|---|---|
| T1 | 0.08 | 0.01 | 0.41 | 0.01 |

| | | | | |
|---|---|---|---|---|
| T2 | 0.11 | 0.04 | 0.25 | 0.04 |
| T3 | 0.06 | 0.03 | 0.14 | 0.02 |

**Supplementary Table S19.** Target sequences for MS2 target identification.

| Strand name | Sequence (5’ → 3’) |
|---|---|
| T1 | ACCACTAATGAGTGATATCC |
| T2 | TACCTGTAGGTAACATGCTC |
| T3 | TCTGCATCCGATTCCATCTC |

**Supplementary Table S20.** 3’ Biotinylated strand sequences for MS2 target identification.

| Strand name | Sequence (5’ → 3’) | Length (nt) |
|---|---|---|
| bT1 | AATGAGTGATATCC/3’-biotin/ | 14 |
| bT2 | TAGGTAACATGCTC/3’-biotin/ | 14 |
| bT3 | TCCGATTCCATCTC/3’-biotin/ | 14 |

**Supplementary Table S21.** Capture strand sequences for MS2 target identification.

| Strand name | Sequence (5’ → 3’) |
|---|---|
| cT1_42 | TTCGACAACTCGTATTAAATCCTTTGCCCGAACGTTATTTTTT GGATATCACTCATTAGTGGT |
| cT2_55 | GTGAGTGAATAACCTTGCTTCTGTAAATCGTCGCTATT TTTTT GAGCATGTTACCTACAGGTA |
| cT3_68 | AGAATATAAAGTACCGACAAAAGGTAAAGTAATTCTGT TTTTT GAGATGGAATCGGATGCAGA |

**Supplementary Table S22.** Guide oligo sequences for MS2 target identification.

| Strand name | Sequence (5’ → 3’) |
|---|---|
| T1a | AACCAACCGAACTGCAACTC |
| T1b | AAGCATCTCATATGCACCCT |
| T2a | GGAGCCAGTCGACAACGAAT |
| T2b | ACGGGGGCCGTAAGGCCCTC |
| T3a | CGATAAGTCTATCGTCGCAA |
| T3b | AACTCCACACCAGGCGATCG |

**Supplementary Table S23.** Complementary strands to MS2 targets with the 30 T-tail used for PAGE gel analysis.

| Strand name | Sequence (5’ → 3’) |
|---|---|
| cM1_30T | TTTTTTTTTTTTTTTTTTTTTTTTTTTTTTGGATATCACTCATTAGTGGT |
| cM2_30T | TTTTTTTTTTTTTTTTTTTTTTTTTTTTTTGAGCATGTTACCTACAGGTA |

### Section S12. Detection of SARS-CoV-2 RNA targets from patient samples using DNA nanobait

Nanobait for SARS-CoV-2 virus is prepared as previously described in Section S2. Below are listed sequences of capture sites (Table S26), biotin strand (Table S25), and target sequence (Table S24). In Table S27 are listed guide oligos for all targets.

**Supplementary Table S24.** Target sequences for SARS-CoV-2 target identification in a patient sample.

| Strand name | Sequence (5' → 3') |
|---|---|
| S1 | CATCCTTACTGCGCTTCGAT |
| S2 | CATTGCAACTGAGGGAGCCT |
| S3 | AGACTCAGACTAATTCTCCT |

**Supplementary Table S25.** 3' Biotinylated strand sequences for SARS-CoV-2 target identification in a patient sample.

| Strand name | Sequence (5' → 3') | Length (nt) |
|---|---|---|
| bS1 | TACTGCGCTTCGAT/3'-biotin/ | 14 |
| bS2 | AACTGAGGGAGCCT/3'-biotin/ | 14 |
| bS3 | AGACTAATTCTCCT/3'-biotin/ | 14 |

**Supplementary Table S26.** Capture strand sequences for SARS-CoV-2 target identification in a patient sample.

| Strand name | Sequence (5' → 3') |
|---|---|
| cS1_43 | TTCGACAACTCGTATTAAATCCTTTGCCCGAACGTTAT TTTTT ATCGAAGCGCAGTAAGGATG |

| | |
|---|---|
| cS2_55 | GTGAGTGAATAACCTTGCTTCTGTAAATCGTCGCTATT TTTTT AGGCTCCCTCAGTTGCAATG |
| cS3_68 | AGAATATAAAGTACCGACAAAAGGTAAAGTAATTCTGT TTTTT AGGAGAATTAGTCTGAGTCT |

**Supplementary Table S27.** Guide DNA oligo sequences for SARS-CoV-2 target identification in patient samples.

| Strand name | Sequence (5’ → 3’) |
|---|---|
| S1a | GCTAGTGTAACTAGCAAGAA |
| S1b | ATTGCAGCAGTACGCACACA |
| S2a | CGTTCTCCATTCTGGTTACT |
| S2b | GTGATCTTTTGGTGTATTCA |
| S3a | GATAACTAGCGCATATACCT |
| S3b | GCTACACTACGTGCCCGCCG |

**Section S13. Detection of SARS-CoV-2 RNA targets from patient samples using DNA nanobait and DNA flower as a label**

The nanobait synthesis follows the previously presented protocol including RNase H cutting and the SDR. As shown in Figure S16, to construct the reference structures on the nanobait, staples 26-32 and 96-102 were substituted by the relevant dumbbell sequences (Table S29). To link the DNA flowers onto the nanobait, staple 43 was substituted by strand P43 and C43 (C43 was added during preparation of 7WJa, so only P43 was added), staple 57 was substituted by strand C57 and P57, and staple 68 was substituted by strand cS3-68. The staples were mixed, and then the linear M13 scaffold was added into the solution (20 nM M13 scaffold, 60 nM staples, 120 nM dumbbell strands, and 120 nM P43, C57, P57, cS3-68). The mixture was heated to 70 °C followed by a linear cooling ramp to 25 °C over 50 minutes. 1 µL of 4 µM 7WJa, 7WJb, and 7WJc (DNA flowers) were added into the 40 µL nanobait solution and incubated at room temperature for 2 h. Finally, the resulted solution was diluted with a washing buffer (10 mM Tris-HCl, 0.5 mM $MgCl_2$, pH 8.0) to 500 µL and centrifuged at 6000 × g for 10 min with an Amicon Ultra 100 kDa filter to remove the excess DNA strands and DNA flowers (repeated 3 times). About 35 µL of nanobait solution was obtained and quantified with a NanoDrop 2000 spectrophotometer.

The flower nanobait for the COVID-19 patient sample was diluted to 250 pM. 3.8 µL of a patient sample (positive or negative samples were processed by the programmable RNase H cutting step) was mixed with 0.2 µL of 250 pM nanobait, 0.5 µL of 100 mM $MgCl_2$, and 0.5 µL of 1 M NaCl. The mixture (5 µL) was incubated at room temperature for 10 min and then diluted by 5 µL of 8 M LiCl and 20 µL of 4 M LiCl (in 1 × TE, pH 9.0) proceeding the nanopore measurement.

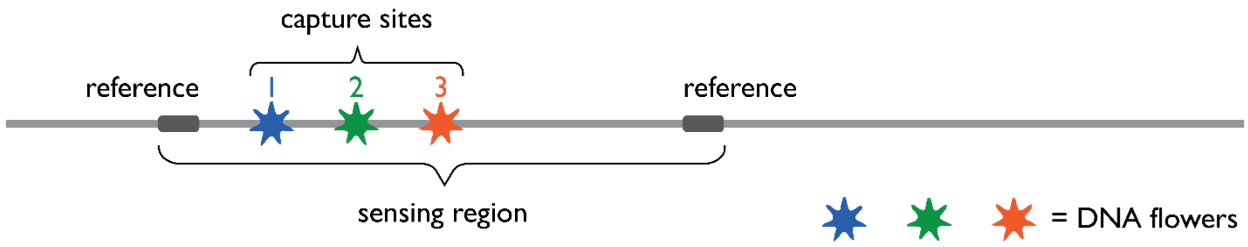

**Figure S16.** Design of nanobait with the DNA flower labels for testing COVID-19 patient sample. Each of the references is represented by eleven closely spaced DNA dumbbells.

Oligonucleotide sequences for DNA flower-based detection with nanobait are listed in Table S28. Example events for negative and positive SARS-CoV-2 patient samples obtained using DNA flowers are shown in Figure S17a and Figure S17b, respectively.

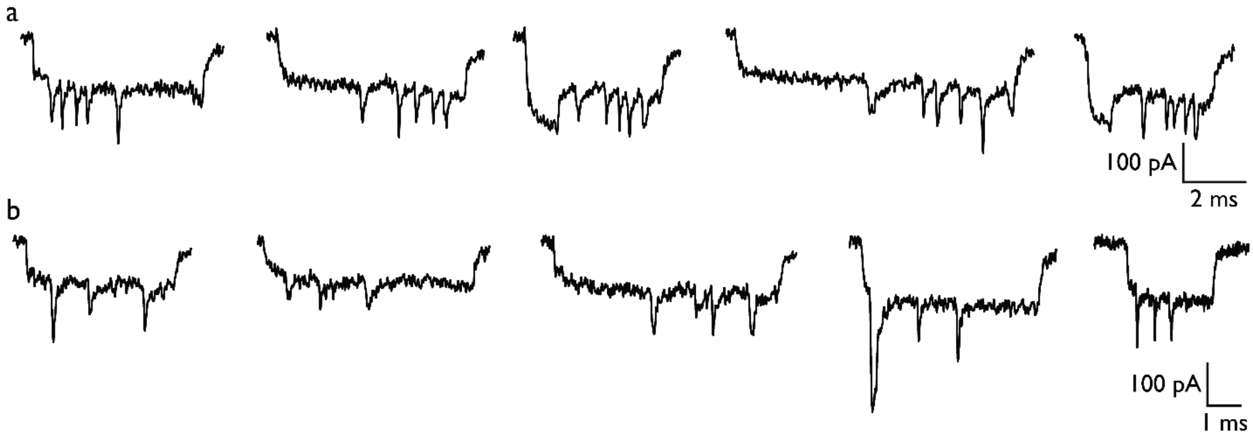

**Figure S17.** Example events of nanobait with DNA flower labels incubated with negative and positive SARS-CoV-2 patient samples (a and b, respectively).

**Supplementary Table S28.** Capture strand sequences for SARS-CoV-2 target identification in a patient sample using DNA flower as a label.

| Strand name | Replaced oligo # | Sequence (5' → 3') |
|---|---|---|
| cFS1_43 | 43 | TAATTTTAAAAGTTTGAGTA TT ATCGAAGCGCAGTA AGGATG |
| 43+ | 43 | ACATTATCATTTTGCGGA |
| cFS2_57 | 57 | AGCTTAGATTAAGACGCTGA TT AGGCTCCCTCAGTT GCAATG |
| 57+ | 57 | GAAGAGTCAATAGTGAAT |
| cFS3_68 | 68 | AGAATATAAAGTACCGACAAAAGGTAAAGTAATTCTGT TTTTT AGGAGAATTAGTCT GAGTCT |

**Supplementary Table S29.** List of oligonucleotides that were replaced from Table S2 to assemble reference structures for nanobait with DNA flower. DNA dumbbell forming sequence is indicated in red.

| Strand name | Sequence (5' → 3') | Length (nt) | Replaced oligo # |
|---|---|---|---|
| *REF 1.1 | CTGAAAGCGTAAGAATACGTGGCACAGACAATATTTTTGAATGGCT | 46 | 26-32 |
| *REF 1.2 | ACATCACTTGTCCTCTTTTGAGGAACAAGTTTTCTTGTCCTGAGTAGA | 48 | |
| *REF 1.3 | AGAACTCAAATCCTCTTTTGAGGAACAAGTTTTCTTGTCTATCGGCCT | 48 | |
| *REF 1.4 | TGCTGGTAATTCCTCTTTTGAGGAACAAGTTTTCTTGTATCCAGAACA | 48 | |
| *REF 1.5 | ATATTACCGCTCCTCTTTTGAGGAACAAGTTTTCTTGTCAGCCATTGC | 48 | |
| *REF 1.6 | AACAGGAAAATCCTCTTTTGAGGAACAAGTTTTCTTGTACGCTCATGG | 48 | |
| *REF 1.7 | AAATACCTACTCCTCTTTTGAGGAACAAGTTTTCTTGTATTTTGACGC | 48 | |
| *REF 1.8 | TCAATCGTCTTCCTCTTTTGAGGAACAAGTTTTCTTGTGAAATGGATT | 48 | |
| *REF 1.9 | ATTTACATTGTCCTCTTTTGAGGAACAAGTTTTCTTGTGCAGATTCAC | 48 | |
| *REF 1.10 | CAGTCACACGTCCTCTTTTGAGGAACAAGTTTTCTTGTACCAGTAATA | 48 | |
| *REF 1.11 | AAAGGGACATTCCTCTTTTGAGGAACAAGTTTTCTTGTTCTGGCCAAC | 48 | |
| *REF 1.12 | AGAGATAGAATCCTCTTTTGAGGAACAAGTTTTCTTGTCCCTTCTGAC | 48 | |
| *REF 2.1 | CTTGAGCCATTCCTCTTTTGAGGAACAAGTTTTCTTGTTTGGGAATTA | 48 | 96-102 |
| *REF 2.2 | GAGCCAGCAATCCTCTTTTGAGGAACAAGTTTTCTTGTAATCACCAGT | 20 | |
| *REF 2.3 | AGCACCATTATCCTCTTTTGAGGAACAAGTTTTCTTGTCCATTAGCAA | 48 | |
| *REF 2.4 | GGCCGGAAACTCCTCTTTTGAGGAACAAGTTTTCTTGTGTCACCAATG | 48 | |
| *REF 2.5 | AAACCATCGATCCTCTTTTGAGGAACAAGTTTTCTTGTTAGCAGCACC | 48 | |
| *REF 2.6 | GTAATCAGTATCCTCTTTTGAGGAACAAGTTTTCTTGTGCGACAGAAT | 48 | |

| | | |
|---|---|---|
| *REF 2.7 | CAAGTTTGCCTCCTCTTTTGAGGAACAAGTTTTCTTGTTTTAGCGTCA | 48 |
| *REF 2.8 | GACTGTAGCGTCCTCTTTTGAGGAACAAGTTTTCTTGTCGTTTTCATC | 48 |
| *REF 2.9 | GGCATTTTCGTCCTCTTTTGAGGAACAAGTTTTCTTGTGTCATAGCCC | 48 |
| *REF 2.10 | CCTTATTAGCTCCTCTTTTGAGGAACAAGTTTTCTTGTGTTTGCCATC | 48 |
| *REF 2.11 | TTTTCATAATTCCTCTTTTGAGGAACAAGTTTTCTTGTCAAAATCACC | 48 |
| *REF 2.12 | GGAACCAGAGCCACCACCGGAACCGCCTCCCTCAGAGCCGCCACCC | 46 |

#### Section S14. Nanopore data analysis

Data analysis of nanopore current traces is performed as previously described [1,4,16]. The data analysis workflow is shown in Figure S18. Firstly, in a raw ionic current trace, our home-built LabView script identifies events by specifying the event duration range and the current threshold. In the next step, nanobait events are separated from isolated events with too small and too large event charge deficit (ECD). Nanobaits can translocate through the nanopore as unfolded or folded (Figure S18). Downward peaks were identified as described previously[1]. The key advantage of nanobait is its pre-determined design relying on the identification of unique current signatures[1,17]. A specific current drop threshold correlating to approximately double the baseline noise was used to identify peaks. Firstly, the reference peaks are identified and positioned. Next, the presence of the peaks corresponding to our sensing sites was identified and an appropriate color was assigned. Lastly, we determine present peaks and calculate displacement efficiency for each site.

Nanopore events can be analyzed with the convolutional neural network QuipuNet following the procedure outlined in Figure S18. Its event detection is much faster, with the rate of 1600 events/s, making it appropriate for instantaneous, real-time data analysis[16] that is not achievable with the other detection methods[18–20].

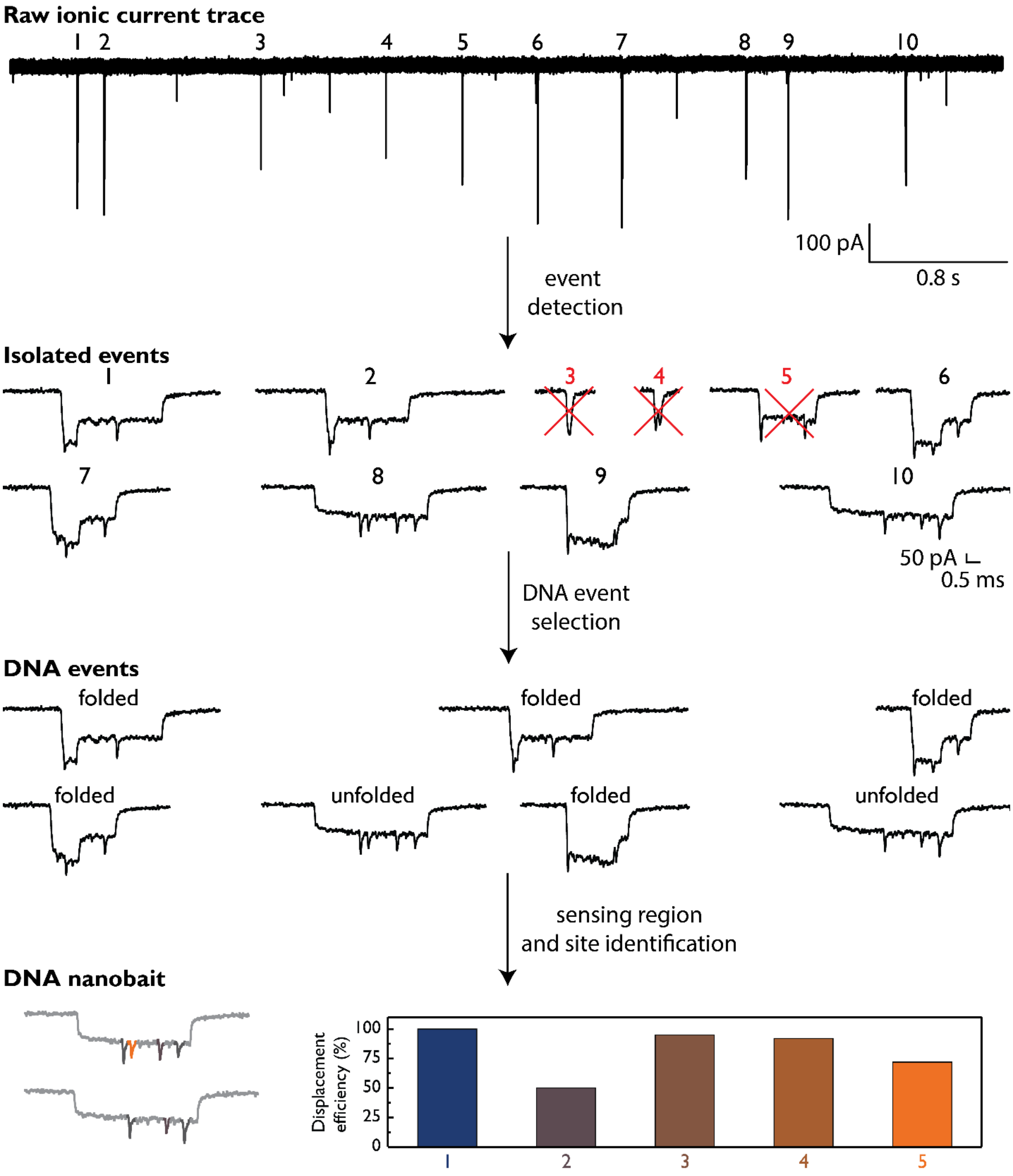

**Figure S18.** Nanopore data analysis workflow. The data analysis starts by searching for single-molecule events from the raw nanopore ionic current traces. Event's duration, current drop, and event charge deficit (ECD i.e. event's surface area) are parameters that further separate nanobait events from DNA events. DNA events in nanopore measurements can include bent or folded events and linear or unfolded events. The next analysis step includes identifying the sensing region (marked by two references) and the absence of the peaks at their respective sites. In the last step, we calculate the mean and errors from multiple single-molecule events (from multiple nanopores), that are subtracted to the control measurement and presented as the displacement efficiency in the bar chart.

### Section S15. Nanopore statistics

Nanopore measurements are listed in Table S30 with characteristics of both nanopores used and sample presented. In all measurements, the first fifty unfolded nanobaits were used for the displacement efficiency calculations unless otherwise specified. All measurements were obtained under an applied field of 600 mV and the respective ionic current value is indicated in Table S30. Only nanopores with the linear current-voltage curve and the noise root mean square (RMS) of < 7 pA were used for nanopore measurements.

**Supplementary Table S30.** Nanopore measurement details for all experiments presented in this study.

| Nanopore measurement | DNA nanobait type / sample name | Current at 600 mV (nA) | Target present | RNase H cutting | Human total RNA | Patient sample | Excess of targets (times) | Incubation time (min) |
|---|---|---|---|---|---|---|---|---|
| 1 | Blank | 10 | / | / | / | / | 10 | 10 |
| 2 | Blank | 11 | / | / | / | / | 10 | 10 |
| 3 | Blank | 14.5 | / | / | / | / | 10 | 10 |
| 4 | Blank | 14 | / | / | / | / | 10 | 10 |
| 5 | Blank | 11 | / | / | / | / | 10 | 10 |
| 6 | Blank | 7 | / | / | / | / | 10 | 10 |
| 7 | Blank | 12 | / | / | / | / | 10 | 10 |
| 8 | Blank | 8 | / | / | / | / | 10 | 10 |
| 9 | H1-H5 | 12.4 | five targets | / | / | / | 10 | 10 |
| 10 | H1-H5 | 9.8 | five targets | / | / | / | 10 | 10 |
| 11 | H1-H5 | 12.15 | five targets | / | / | / | 10 | 10 |
| 12 | H1-H5 | 12.3 | five targets | / | / | / | 10 | 10 |
| 13 | total RNA + Nanobait | 9.5 | / | / | yes | / | 10 | 10 |
| 14 | total RNA + Nanobait | 10.2 | / | / | yes | / | 10 | 10 |
| 15 | total RNA + Nanobait | 10 | / | / | yes | / | 10 | 10 |
| 16 | total RNA + Nanobait+targets | 9.4 | five targets | / | yes | / | 10 | 10 |
| 17 | total RNA + Nanobait+targets | 9.25 | five targets | / | yes | / | 10 | 10 |
| 18 | total RNA + Nanobait+targets | 10.5 | five targets | / | yes | / | 10 | 10 |

| | | | | | | | | |
|---|---|---|---|---|---|---|---|---|
| 19 | Nanobait + negative covid sample a53 | 9 | / | / | / | negative | NA | 10 |
| 20 | Nanobait + negative covid sample a53 | 9 | / | / | / | negative | NA | 10 |
| 21 | Nanobait + negative covid sample a53 | 10.6 | / | / | / | negative | NA | 10 |
| 22 | Nanobait + negative covid sample a55 | 10.2 | / | / | / | negative | NA | 10 |
| 23 | Nanobait + negative covid sample a55 | 11 | / | / | / | negative | NA | 10 |
| 24 | Nanobait + negative covid sample a55 | 8.72 | / | / | / | negative | NA | 10 |
| 25 | Nanobait + negative covid sample+targets | 10.5 | / | / | / | negative | NA | 10 |
| 26 | Nanobait + negative covid sample+targets | 10.25 | / | / | / | negative | NA | 10 |
| 27 | Nanobait + negative covid sample+targets | 11 | / | / | / | negative | NA | 10 |
| 28 | Nanobait + negative covid sample+RNaseH cutting | 10.2 | / | yes | / | negative | NA | 10 |
| 29 | Nanobait + negative covid sample+RNaseH cutting | 9.8 | / | yes | / | negative | NA | 10 |
| 30 | kinetics 1 min DNA | 9.8 | all targets | / | / | / | 10 | 1 |
| 31 | kinetics 2.5 min DNA | 10.9 | all targets | / | / | / | 10 | 2.5 |
| 32 | kinetics 5 min DNA | 11.2 | all targets | / | / | / | 10 | 5 |
| 33 | kinetics 10 min DNA | 10 | all targets | / | / | / | 10 | 10 |
| 34 | kinetics 1 min RNA | 12 | all targets | / | / | / | 10 | 1 |
| 35 | kinetics 2.5 min RNA | 12.4 | all targets | / | / | / | 10 | 2.5 |
| 36 | kinetics 5 min RNA | 12.2 | all targets | / | / | / | 10 | 5 |
| 37 | kinetics 10 min RNA | 10 | all targets | / | / | / | 10 | 10 |
| 38 | Nanobait for MS2 blank | 12 | / | / | / | / | 10 | 10 |
| 39 | Nanobait for MS2 blank | 11 | / | / | / | / | 10 | 10 |
| 40 | Nanobait for MS2 blank | 11 | / | / | / | / | 10 | 10 |
| 41 | Nanobait for MS2 blank+targets | 11.58 | all targets | / | / | / | 10 | 10 |
| 42 | Nanobait for MS2 blank+targets | 10.8 | all targets | / | / | / | 10 | 10 |

| | | | | | | | | |
|---|---|---|---|---|---|---|---|---|
| 43 | Nanobait for MS2 blank+RNaseH cutting | 10 | / | yes | / | / | 10 | 10 |
| 44 | Nanobait for MS2 blank+RNaseH cutting | 12.5 | / | yes | / | / | 10 | 10 |
| 45 | Nanobait for MS2 blank+RNaseH cutting | 13.1 | / | yes | / | / | 10 | 10 |
| 46 | Nanobait for a patient covid sample | 11 | / | / | / | / | 10 | 10 |
| 47 | Nanobait for a patient covid sample | 10.7 | / | / | / | / | 10 | 10 |
| 48 | Nanobait for patient covid sample+negative sample | 9.9 | / | yes | / | negative | NA | 10 |
| 49 | Nanobait for patient covid sample+positive sample | 9.8 | / | yes | / | positive | NA | 10 |
| 50 | Nanobait for patient covid sample+positive sample | 10.7 | / | yes | / | positive | NA | 10 |
| 51 | Nanobait for patient covid sample+positive sample | 10.4 | / | yes | / | positive | NA | 10 |
| 52 | Nanobait for patient covid sample with DNA flower | 6.98 | / | / | / | / | 10 | 10 |
| 53 | Nanobait for patient covid sample with DNA flower+negative sample | 11.4 | / | yes | / | negative | NA | 10 |
| 54 | Nanobait for patient covid sample with DNA flower+negative sample | 11.18 | / | yes | / | negative | NA | 10 |
| 55 | Nanobait for patient covid sample with DNA flower | 11.44 | / | yes | / | positive | NA | 10 |
| 56 | Nanobait for patient covid sample with DNA flower | 9.89 | / | yes | / | positive | NA | 10 |
| 57 | Nanobait for patient covid sample with DNA flower | 10.1 | / | yes | / | positive | NA | 10 |
| 58 | Nanobait for multiple viruses blank | 9 | / | / | / | / | 10 | 10 |

| | | | | | | | | |
|---|---|---|---|---|---|---|---|---|
| 59 | Nanobait for multiple viruses blank | 7.5 | / | / | / | / | 10 | 10 |
| 60 | Nanobait for multiple viruses blank | 8.5 | / | / | / | / | 10 | 10 |
| 61 | Nanobait for multiple viruses blank+all targets | 8.8 | all targets | / | / | / | 10 | 10 |
| 62 | Nanobait for multiple viruses blank+all targets | 8 | all targets | / | / | / | 10 | 10 |
| 63 | Nanobait for multiple viruses blank+all targets | 11 | all targets | / | / | / | 10 | 10 |
| 64 | Nanobait for multiple viruses blank+SARS-CoV-2 target | 8.6 | SARS-CoV-2 | / | / | / | 10 | 10 |
| 65 | Nanobait for multiple viruses blank+SARS-CoV-2 target | 10.15 | SARS-CoV-2 | / | / | / | 10 | 10 |
| 66 | Nanobait for multiple viruses blank+SARS-CoV-2 target | 11.4 | SARS-CoV-2 | / | / | / | 10 | 10 |
| 67 | Nanobait for multiple viruses blank+Influenza target | 8.99 | Influenza | / | / | / | 10 | 10 |
| 68 | Nanobait for multiple viruses blank+Influenza target | 9.42 | Influenza | / | / | / | 10 | 10 |
| 69 | Nanobait for multiple viruses blank+Influenza target | 14.8 | Influenza | / | / | / | 10 | 10 |
| 70 | Nanobait for multiple viruses blank+RSV target | 9.54 | RSV | / | / | / | 10 | 10 |
| 71 | Nanobait for multiple viruses blank+RSV target | 15.15 | RSV | / | / | / | 10 | 10 |
| 72 | Nanobait for multiple viruses blank+RSV target | 9.3 | RSV | / | / | / | 10 | 10 |
| 73 | Nanobait for multiple viruses blank+Parainfluenza target | 9.6 | Parainfluenza | / | / | / | 10 | 10 |
| 74 | Nanobait for multiple viruses | 9.67 | Parainfluenza | / | / | / | 10 | 10 |

| | | | | | | | | |
|---|---|---|---|---|---|---|---|---|
| | blank+Parainfluenza target | | | | | | | |
| 75 | Nanobait for multiple viruses blank+Parainfluenza target | 15.2 | Parainfluenza | / | / | / | 10 | 10 |
| 76 | Nanobait for multiple viruses blank+Rhinoviruses target | 8.9 | Rhinoviruses | / | / | / | 10 | 10 |
| 77 | Nanobait for multiple viruses blank+Rhinoviruses target | 15.5 | Rhinoviruses | / | / | / | 10 | 10 |
| 78 | Nanobait for multiple viruses blank+Rhinoviruses target | 9.74 | Rhinoviruses | / | / | / | 10 | 10 |
| 79 | Nanobait for variants blank | 9.01 | / | / | / | / | 10 | 10 |
| 80 | Nanobait for variants blank | 9.7 | / | / | / | / | 10 | 10 |
| 81 | Nanobait for variants blank | 8.52 | / | / | / | / | 10 | 10 |
| 82 | Nanobait for variants blank + WT targets | 8.5 | all targets | / | / | / | 10 | 10 |
| 83 | Nanobait for variants blank + WT targets | 8.5 | all targets | / | / | / | 10 | 10 |
| 84 | Nanobait for variants blank + WT targets | 7.8 | all targets | / | / | / | 10 | 10 |
| 85 | Nanobait for variants blank + variant targets | 8.8 | all targets | / | / | / | 10 | 10 |
| 86 | Nanobait for variants blank + variant targets | 9.03 | all targets | / | / | / | 10 | 10 |
| 87 | Nanobait for variants blank + variant targets | 9 | all targets | / | / | / | 10 | 10 |
| 88 | Nanobait for variants+SARS-CoV-2 reference variant target | 11.5 | SARS-CoV-2 reference | / | / | / | 10 | 10 |
| 89 | Nanobait for variants+ SARS-CoV-2 reference variant target | 10.5 | SARS-CoV-2 reference | / | / | / | 10 | 10 |
| 90 | Nanobait for variants+ SARS-CoV-2 reference variant target | 9.7 | SARS-CoV-2 reference | / | / | / | 10 | 10 |

| | | | | | | | | |
|---|---|---|---|---|---|---|---|---|
| 91 | Nanobait for variants+Delta variant target | 10 | Delta | / | / | / | 10 | 10 |
| 92 | Nanobait for variants+Delta variant target | 10 | Delta | / | / | / | 10 | 10 |
| 93 | Nanobait for variants+Delta variant target | 12 | Delta | / | / | / | 10 | 10 |
| 94 | Nanobait for variants+ B.1 variant target | 10.1 | B.1 | / | / | / | 10 | 10 |
| 95 | Nanobait for variants+ B.1 variant target | 13.75 | B.1 | / | / | / | 10 | 10 |
| 96 | Nanobait for variants+ B.1 variant target | 12.8 | B.1 | / | / | / | 10 | 10 |
| 97 | Nanobait for variants+Alpha variant target | 10.68 | Alpha | / | / | / | 10 | 10 |
| 98 | Nanobait for variants+Alpha variant target | 8.3 | Alpha | / | / | / | 10 | 10 |
| 99 | Nanobait for variants+Alpha variant target | 11.3 | Alpha | / | / | / | 10 | 10 |
| 100 | Nanobait for variants+Beta variant target | 9.5 | Beta | / | / | / | 10 | 10 |
| 101 | Nanobait for variants+Beta variant target | 9.7 | Beta | / | / | / | 10 | 10 |
| 102 | Nanobait for variants+Beta variant target | 14.1 | Beta | / | / | / | 10 | 10 |
| 103 | Nanobait for patient covid sample+negative sample | 11 | / | yes | / | negative | NA | 10 |
| 104 | Nanobait for patient covid sample+negative sample | 12 | / | yes | / | negative | NA | 10 |
| 105 | Nanobait for patient covid sample+negative sample | 9.7 | / | yes | / | negative | NA | 10 |
| 106 | Nanobait for patient covid sample+negative sample | 11 | / | yes | / | negative | NA | 10 |
| 107 | Nanobait for patient covid | 11.5 | / | yes | / | negative | NA | 10 |

| | | | | | | | | |
|---|---|---|---|---|---|---|---|---|
| | sample+negative sample | | | | | | | |
| 108 | Nanobait for patient covid sample+negative sample | 10 | / | yes | / | negative | NA | 10 |
| 109 | Nanobait for patient covid sample+negative sample | 11.1 | / | yes | / | negative | NA | 10 |
| 110 | Nanobait for patient covid sample+negative sample | 11.3 | / | yes | / | negative | NA | 10 |
| 111 | Nanobait for patient covid sample+negative sample | 11.8 | / | yes | / | negative | NA | 10 |
| 112 | Nanobait for SARS-CoV-2 RNA N501 | 7 | / | / | / | / | 10 | 10 |
| 113 | Nanobait for SARS-CoV-2 RNA N501+wild-type | 8.36 | WT | yes | / | / | 10 | 10 |
| 114 | Nanobait for SARS-CoV-2 RNA N501+wild-type | 7.6 | WT | yes | / | / | 10 | 10 |
| 115 | Nanobait for SARS-CoV-2 RNA N501+wild-type | 5.6 | WT | yes | / | / | 10 | 10 |
| 116 | Nanobait for SARS-CoV-2 RNA N501+N501S | 12.78 | N501S | yes | / | / | 10 | 10 |
| 117 | Nanobait for SARS-CoV-2 RNA N501+ N501S | 8.7 | N501S | yes | / | / | 10 | 10 |
| 118 | Nanobait for SARS-CoV-2 RNA N501+ N501S | 9 | N501S | yes | / | / | 10 | 10 |
| 119 | Nanobait for SARS-CoV-2 RNA N501+N501T | 12.6 | N501T | yes | / | / | 10 | 10 |
| 120 | Nanobait for SARS-CoV-2 RNA N501+ N501T | 8.2 | N501T | yes | / | / | 10 | 10 |
| 121 | Nanobait for SARS-CoV-2 RNA N501+ N501T | 14 | N501T | yes | / | / | 10 | 10 |

#### Section S16. Sensitivity curve

We prepared serial dilutions of nanobait with SARS-CoV-2 N501 WT cut RNA (Figure S19a; details in Section S8). Any contact surfaces were passivated with a 20 bp DNA (5 nM; 5'-GACCACTACAGTTGTAATCC-3' IDT) prior to contact with the diluted samples to prevent surface binding. The sensitivity curve was shown from fM to nM range of nanobait concentrations.

Nanobait was mixed with SARS-CoV-2 RNA targets (20 nt, H2 and H5 sequences in Supplementary Table S15) in various ratios of RNA for 10 min and by keeping nanobait concentration constant (Figure S19b).

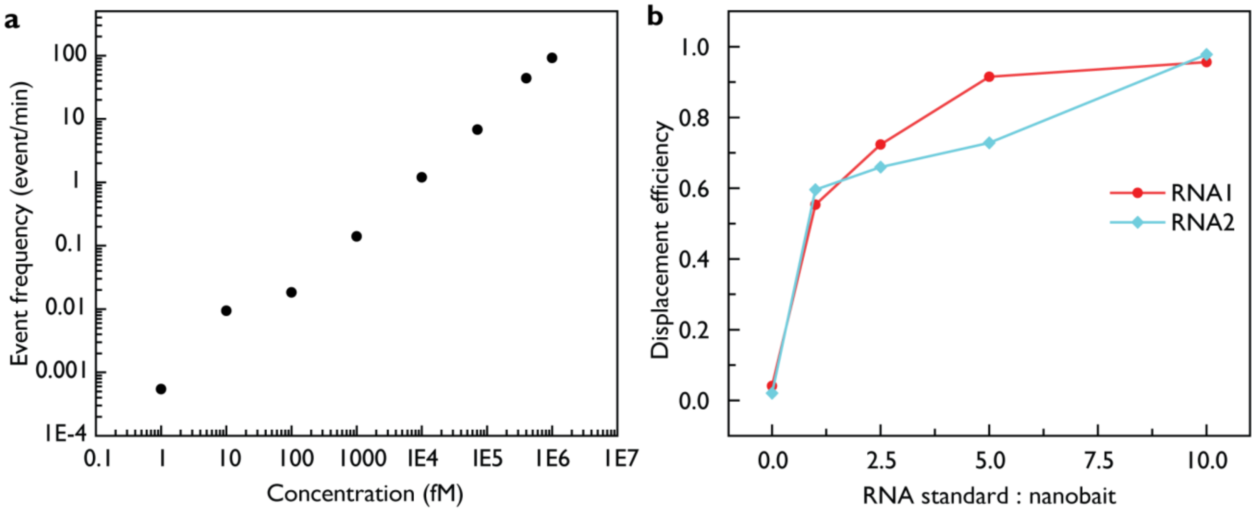

**Figure S19.** Sensitivity curve of nanobait-nanopore system. **a)** Nanobait detection with SARS-CoV-2 N501 wild-type target as described in Section S8. Were detected in the range from femtomolar (fM) to nanomolar (nM) concentrations. Event frequency was plotted as a function of log of nanobait concentration in fM versus log of event frequency in events per min. **b)** Nanobait displacement efficiency with variable SARS-CoV-2 RNA concentrations. Nanobait concentration was constant at 500 pM and viral RNAs (RNA1 and RNA2 correspond to H2 and H4, respectively; 20 nt) were varied from 1:1 ratio to 1:10 excess to nanobait to test dynamic range of nanobait.
